# Epidemiologic transition of lung cancer mortality in Italy by sex, province of residence and birth cohort (1920-1929 to 1960-1969)

**DOI:** 10.1101/2023.01.26.23285051

**Authors:** Dolores Catelan, Annibale Biggeri, Lauro Bucchi, Valerio Manno, Marilena Pappagallo, Giorgia Stoppa, Francesco Grippo, Luisa Frova, Federica Zamagni, Roberta Crialesi, Giada Minelli

**Author notes:** **Correspondence** Federica Zamagni, Emilia-Romagna Cancer Registry, Romagna Cancer Institute, IRCCS Istituto Romagnolo per lo Studio dei Tumori (IRST) Dino Amadori, 47014 Meldola, Forlì, Italy. **Funding information** This study did not receive any external funding.

## Abstract

Space-time analysis of mortality risk is useful to evaluate the epidemiologic transitions at the subnational level. In this study, we analysed the death certificate records for lung cancer in Italy in 1995-2016, obtained from the Italian National Statistics Institute. Our objective was to investigate the spatio-temporal evolution of lung cancer mortality by sex and province of residence (n = 107) using the birth cohort as relevant time axis. We built space-time Bayesian models with space-time interactions. Among men (n = 554,829), mortality peaked in the 1920-1929 cohort, followed by a generalised decline. Among women (n = 158,619), we found novel original evidence for a peak in the 1955-1964 cohort, equivalent to a 35-year delay, with a downward trend being observed thereafter. Over time, the documented north-south decreasing mortality gradient has been replaced by a west-east decreasing gradient. Naples has become the province at highest risk in Italy, both among men and women. This pattern is consistent with an epidemiologic transition of risk factors for lung cancer to the south-west of the country and raises concerns, because 5-year age-standardised net survival from the disease in this geographic area is lower than in northern and central Italy. The variability of mortality rates among provinces has changed over time, with an increasing homogeneity for men and an opposite trend for women in the more recent birth cohorts. These unprecedented observations update substantially previous knowledge on lung cancer mortality in Italy.

**What’s new?:** The epidemiologic transition of lung cancer mortality in Italy (1995-2016) was studied using space-time Bayesian models with space-time interactions. Among men, mortality peaked in the 1920-1929 cohort, followed by a decline. Among women, novel evidence was found for a peak in the 1955-1964 cohort, equivalent to a 35-year delay, with a downward trend thereafter. The north-south decreasing gradient has been replaced by a west-east decreasing gradient, with Naples currently being the province at highest risk.

## 1 INTRODUCTION

Lung cancer mortality trends in space and time across Europe reflect the geography and evolution of the smoking epidemic and, secondarily, of exposure to nontobacco risk factors.^1^ Their knowledge may offer insights into future perspectives for lung cancer control in every single country and region.^2^ At present, in fact, there are no consistent geographic patterns of lung cancer mortality on a continental scale. For men, the highest rates are encountered, for example, in Poland and some French areas, whereas, among women, peaks are also observed in Germany and Iceland.^3^ Cross-sectional comparisons, however, are unwarranted, because different countries may be in different phases of the smoking/lung cancer epidemic^4^ and of their economic trajectory, and these phases may be years or decades apart from each other. Differences concerning the intensity of tobacco control programmes may further flaw international cross-sectional mortality comparisons.^4-6^

Contrasting country-specific time trends in mortality is more informative of the evolution of the epidemic. Among men, mortality rates began to decrease in the late 1970s in Sweden, Finland and the U.K. and during the 1980s in The Netherlands, Belgium and Switzerland, followed by Spain and Eastern Europe in the 1990s.^6^ Underlying these trends were birth-cohort-dependent changes in the risk of lung cancer death. In Sweden, Finland, and the U.K., mortality dropped starting with generations born in the first decade of the past century. In southern Europe, a decrease occurred only for cohorts born in the middle of the century and after.^6^

Regarding women, the spatial-temporal pattern of lung cancer mortality is more complex. During the 1980s, the rates plateaued in the U.K., Ireland and Iceland but not in Finland, Sweden and Norway. In western Europe, mortality is still rising, particularly in France and The Netherlands. During the 1990s, the rates stabilised in some, but not all, eastern European countries. In most southern European countries, mortality is lower but still increasing.^6^ Importantly, in the few countries where female mortality has begun to decrease, the downturn has occurred some years or decades later than in the male population.^6,7^

In Italy, the average lung cancer mortality rates over the past two decades ranked slightly below the European median in both sexes.^5^ The annual change in mortality had a similar ranking position for men as for women, but in opposite directions. Like in Spain and Greece, the trend declined around 1990 for men, while continuing to increase for the female population.^8^ Around 2010, however, some reports provided an early indication that mortality was probably reaching a peak among younger women.^9,10^ The stable trend in incidence observed since 2003 among women aged <50 years was consistent with this hypothesis.^11^ So far, however, no sound evidence has been presented for a stable reduction in lung cancer risk for women in the most recent generations. Also, no research has been published on whether the evolution of the epidemic by sex differs geographically within Italy. Incidentally, internal national geographic differences in lung cancer mortality trends have seldom been explored both in the European countries^12^ and elsewhere.^13^

The above considerations provided the rationale for conducting an updated analysis of the epidemiologic transition of lung cancer in Italy. We studied the mortality trend in 1995-2016, by sex, birth cohort, and province of residence.

## 2 MATERIALS AND METHODS

### 2.1 Rationale and design

The temporal evolution of mortality can be analysed along two different time axes, i.e. the birth cohort or the calendar period. Changes in the prevalence of risk factors for lung cancer usually have greater effects on the former.^4,14^ However, the most common approach to the analysis of the temporal evolution of lung cancer is a period approach, because the analysis by birth cohort requires the availability of data for a long time span.

Space-time variation in mortality is particularly suited to evaluate the evolution of the mortality gradient across geographic areas and the presence of migration of risk factors. Researchers have generally considered large-scale data collected at a national^15^ or international level.^4-6^ Two examples of analysis on a small geographic scale are in studies by Dreassi et al.^16^ and Catelan et al.,^17^ which dealt with the space-time variation in lung cancer mortality at the municipality level in the male population of an Italian administrative region. The period and, respectively, the birth cohort were considered relevant time axes. An example of bivariate disease mapping, with disease mortality being studied in the same area in both sexes through Bayesian space-time models, can be found in Biggeri et al.^9^

In the present study, we evaluated the temporal and spatial evolution of lung cancer mortality by sex and province of residence, taking the birth cohort as a relevant time axis. The analysis was performed using a Bayesian space-time model in which space-time interaction was allowed.

### 2.2 Mortality data

The Italian National Institute of Statistics (ISTAT) and the Italian National Institute of Health (ISS) made available to us the anonymous records of lung cancer death certificates of all Italian men (n = 554,829) and women (n = 158,619) who died between 1995 and 2016. Records were extracted by the Statistical Service of ISS from the ISTAT cause-specific mortality database using the codes for lung cancer from the International Classification of Diseases (ICD), i.e. 162 (according to the ICD-9)^18^ from 1995 to 2002 and C33-C34 (ICD-10)^19^ thereafter.

The provinces of residence (n = 107) were defined according to the 2016 criteria from the Italian National Institute of Statistics and were identified by grouping the recorded municipalities. For each province, residents and deaths were cross-classified into 18 age groups (0-4, …, 85+) and five calendar periods (1995-1999, …, 2015-2016).

For the space-time analysis, we considered nine birth cohorts (1920-1929, …, 1960-1969) corresponding to people aged 30-74 years at the beginning of the study period. We focused on these cohorts because they were followed-up for the whole period and yielded a substantial number of events. Supplementary Figure 1 shows the correspondence between birth cohorts, age groups and calendar periods.

We fitted age-period-cohort models on the data for the whole of Italy to estimate adjusted mortality rate ratios for calendar periods and birth cohorts. For men, there were large death counts and log-linear models with many parameters tended to be well-supported. This is a known phenomenon in the analysis of contingency tables.^20^ To be parsimonious, we opted for a forward selection strategy starting with the simplest model and testing for adding further parameters while adjusting for overdispersion. For women, data counts were on average 4.7 times smaller. However, we did not find any overdispersion and the forward or backward selection strategies gave the same results.

We also computed directly standardised mortality rates using the 2013 revision of the European standard population.^21^ On average, the rates standardised in this way are 60% higher than they would be if the 1976 European standard population was used. Standardised mortality ratios (SMRs) were computed using an internal indirect standardisation.^22^

For the analysis by province, the expected number of deaths was calculated using age-specific reference rates derived from the age-cohort model fitted at the national level.^23^ In this way, the expected number of cases was not influenced by cohort effects, which could then be specified as parameters in the Bayesian models. We assumed the multiplicative age-cohort model^24^ to be valid. Other studies have shown examples in which the interaction terms or period effects are not relevant.^25,26^ Observed and expected cases were aggregated along the diagonals of the Lexis diagram representing the abovementioned nine birth cohorts, thus collapsing over the age dimension by province.

### 2.3 The space-cohort Bayesian model

We described the overall evolution in space and time of lung cancer mortality in Italy through a space-time model in which birth cohort was taken as the relevant time axis. Let us assume that O_i,j_, the number of observed cases in the i-th province (i = 1, …,107) for the j-th birth cohort (j = 1920-1929, …, 1960-1969), follows a Poisson distribution with mean E_i,j_θ_i,j_, where E_i,j_ indicates the expected number of cases under indirect standardisation and θ_i,j_ the relative risk (RR). Following the specification of Besag et al.^27^ a random effects model is assumed for the logarithm of RR

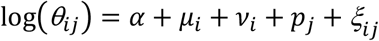

where *α* represents the intercept, for which an *a priori* improper uniform is assumed; *μ*_*i*_ a spatially structured random term and *ν*_*i*_ a spatially unstructured random term. The term *μ*_*i*_, called clustering random term, captures the Poisson overdispersion, which is spatially structured and shrinks the relative risk towards a local mean. The clustering component *μ*_*i*_ is modelled, conditionally on *μ*_*l*∼*i*_ terms (*l∼i* denotes the set of adjacent areas to the *i*-th one, index *l* assumes all integers from *1* to *n*_*i*_, the number of adjacent areas to the *i*-th one) as Normal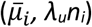 where:

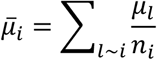

The term *ν*_*i*_, called the heterogeneity random term, captures the Poisson overdispersion, which is not spatially structured and stabilises the relative risk toward the global mean. The *a priori* distribution for the heterogeneity is assumed to be Normal(*0, λ*_*v*_). The hyperprior distributions of the precision parameters *λ*_*v*_, *λ*_*u*_ are assumed to be Gamma (*0*.*5,0*.*0005*).^28^ The random term *p*_*j*_ represents the effect of the j-th cohort whose a priori distribution is assumed to be a first-order random walk with normal independent increments. The term *ξ*_*ij*_ represents the space-time interaction, whose prior distribution can be specified in several ways depending on the assumptions about the dependence structure. In our model, we assumed that the interaction terms are structured both in space and time^29^ (see Dreassi et al. for methodological details).^16^

## 3 Results

### 3.1 Age and sex specific time evolution of lung cancer mortality rates

Between 1995 and 2016, 554,829 deaths were observed among men and 158,619 among women, for an age-standardised mortality rate of 101.7 per 100,000 and 21.5 per 100,000, respectively. Table 1 shows the 5-year age-specific mortality rates and death counts by 5-year calendar period and sex. The rates by column represent the cross-sectional age-specific curve of mortality rates for each calendar period. The rates by diagonal represent the longitudinal age-specific curve of mortality rates of each birth cohort. In Supplementary Figure 1, the birth cohorts corresponding to the diagonals of the Lexis diagram are shown. Highlighted in grey are the birth cohorts used for the spatio-temporal analysis. In Figure 1, the rates by columns represent the cross-sectional age-specific curve of mortality rates for each calendar period. The rates by diagonals represent the longitudinal age-specific curve of mortality rates of each birth cohort.

**TABLE 1.**
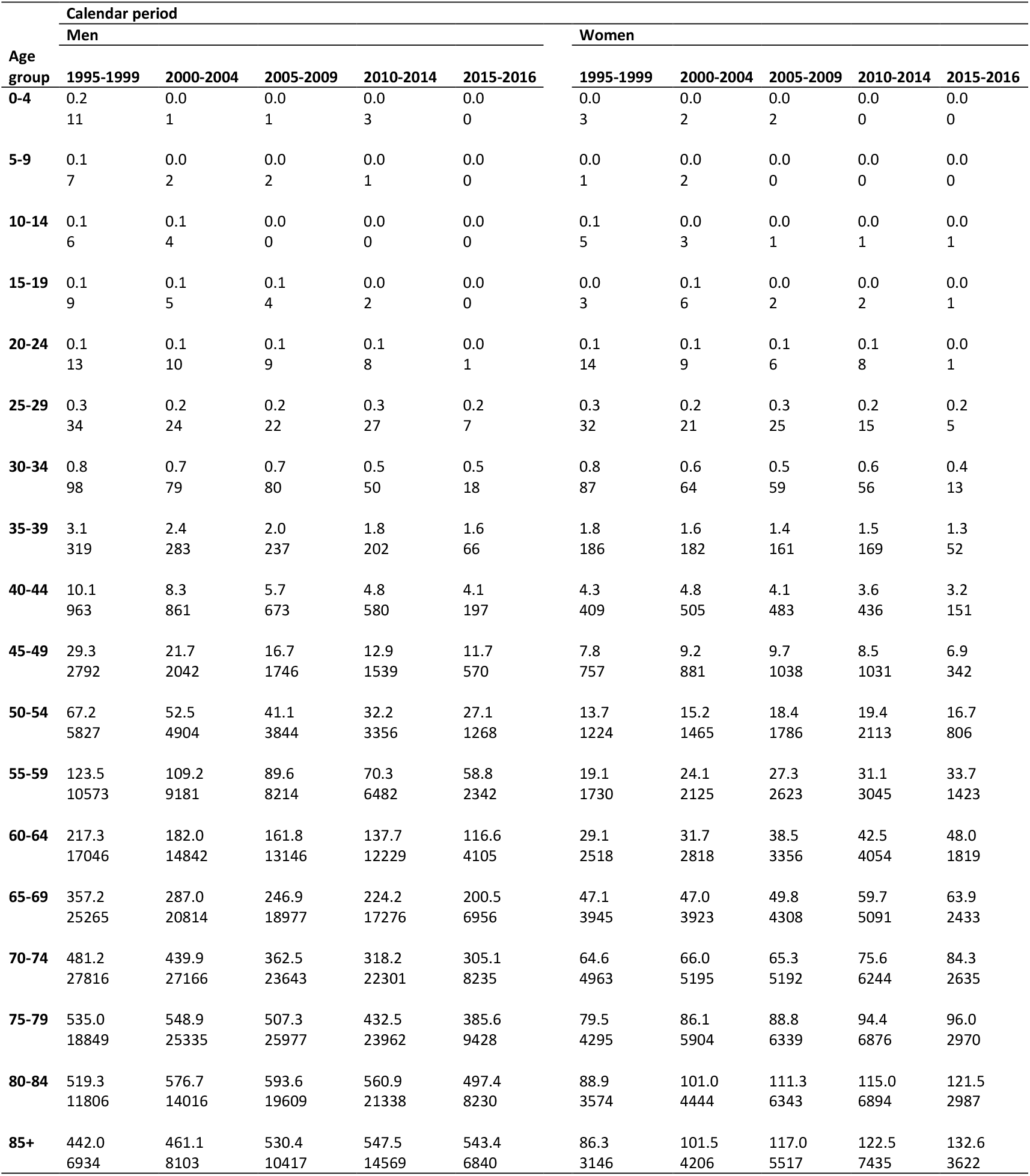
Observed lung cancer mortality rates per 100,000 person-years and number of deaths by sex, calendar period, and age group. Italy, 1995-2016

| Age group | Calendar period |  |  |  |  |  |  |  |  |  |
| --- | --- | --- | --- | --- | --- | --- | --- | --- | --- | --- |
|  | Men |  |  |  |  | Women |  |  |  |  |
|  | 1995-1999 | 2000-2004 | 2005-2009 | 2010-2014 | 2015-2016 | 1995-1999 | 2000-2004 | 2005-2009 | 2010-2014 | 2015-2016 |
| **0-4** | 0.2<br>11 | 0.0<br>1 | 0.0<br>1 | 0.0<br>3 | 0.0<br>0 | 0.0<br>3 | 0.0<br>2 | 0.0<br>2 | 0.0<br>0 | 0.0<br>0 |
| **5-9** | 0.1<br>7 | 0.0<br>2 | 0.0<br>2 | 0.0<br>1 | 0.0<br>0 | 0.0<br>1 | 0.0<br>2 | 0.0<br>0 | 0.0<br>0 | 0.0<br>0 |
| **10-14** | 0.1<br>6 | 0.1<br>4 | 0.0<br>0 | 0.0<br>0 | 0.0<br>0 | 0.1<br>5 | 0.0<br>3 | 0.0<br>1 | 0.0<br>1 | 0.0<br>1 |
| **15-19** | 0.1<br>9 | 0.1<br>5 | 0.1<br>4 | 0.0<br>2 | 0.0<br>0 | 0.0<br>3 | 0.1<br>6 | 0.0<br>2 | 0.0<br>2 | 0.0<br>1 |
| **20-24** | 0.1<br>13 | 0.1<br>10 | 0.1<br>9 | 0.1<br>8 | 0.0<br>1 | 0.1<br>14 | 0.1<br>9 | 0.1<br>6 | 0.1<br>8 | 0.0<br>1 |
| **25-29** | 0.3<br>34 | 0.2<br>24 | 0.2<br>22 | 0.3<br>27 | 0.2<br>7 | 0.3<br>32 | 0.2<br>21 | 0.3<br>25 | 0.2<br>15 | 0.2<br>5 |
| **30-34** | 0.8<br>98 | 0.7<br>79 | 0.7<br>80 | 0.5<br>50 | 0.5<br>18 | 0.8<br>87 | 0.6<br>64 | 0.5<br>59 | 0.6<br>56 | 0.4<br>13 |
| **35-39** | 3.1<br>319 | 2.4<br>283 | 2.0<br>237 | 1.8<br>202 | 1.6<br>66 | 1.8<br>186 | 1.6<br>182 | 1.4<br>161 | 1.5<br>169 | 1.3<br>52 |
| **40-44** | 10.1<br>963 | 8.3<br>861 | 5.7<br>673 | 4.8<br>580 | 4.1<br>197 | 4.3<br>409 | 4.8<br>505 | 4.1<br>483 | 3.6<br>436 | 3.2<br>151 |
| **45-49** | 29.3<br>2792 | 21.7<br>2042 | 16.7<br>1746 | 12.9<br>1539 | 11.7<br>570 | 7.8<br>757 | 9.2<br>881 | 9.7<br>1038 | 8.5<br>1031 | 6.9<br>342 |
| **50-54** | 67.2<br>5827 | 52.5<br>4904 | 41.1<br>3844 | 32.2<br>3356 | 27.1<br>1268 | 13.7<br>1224 | 15.2<br>1465 | 18.4<br>1786 | 19.4<br>2113 | 16.7<br>806 |
| **55-59** | 123.5<br>10573 | 109.2<br>9181 | 89.6<br>8214 | 70.3<br>6482 | 58.8<br>2342 | 19.1<br>1730 | 24.1<br>2125 | 27.3<br>2623 | 31.1<br>3045 | 33.7<br>1423 |
| **60-64** | 217.3<br>17046 | 182.0<br>14842 | 161.8<br>13146 | 137.7<br>12229 | 116.6<br>4105 | 29.1<br>2518 | 31.7<br>2818 | 38.5<br>3356 | 42.5<br>4054 | 48.0<br>1819 |
| **65-69** | 357.2<br>25265 | 287.0<br>20814 | 246.9<br>18977 | 224.2<br>17276 | 200.5<br>6956 | 47.1<br>3945 | 47.0<br>3923 | 49.8<br>4308 | 59.7<br>5091 | 63.9<br>2433 |
| **70-74** | 481.2<br>27816 | 439.9<br>27166 | 362.5<br>23643 | 318.2<br>22301 | 305.1<br>8235 | 64.6<br>4963 | 66.0<br>5195 | 65.3<br>5192 | 75.6<br>6244 | 84.3<br>2635 |
| **75-79** | 535.0<br>18849 | 548.9<br>25335 | 507.3<br>25977 | 432.5<br>23962 | 385.6<br>9428 | 79.5<br>4295 | 86.1<br>5904 | 88.8<br>6339 | 94.4<br>6876 | 96.0<br>2970 |
| **80-84** | 519.3<br>11806 | 576.7<br>14016 | 593.6<br>19609 | 560.9<br>21338 | 497.4<br>8230 | 88.9<br>3574 | 101.0<br>4444 | 111.3<br>6343 | 115.0<br>6894 | 121.5<br>2987 |
| **85+** | 442.0<br>6934 | 461.1<br>8103 | 530.4<br>10417 | 547.5<br>14569 | 543.4<br>6840 | 86.3<br>3146 | 101.5<br>4206 | 117.0<br>5517 | 122.5<br>7435 | 132.6<br>3622 |

**FIGURE 1.**
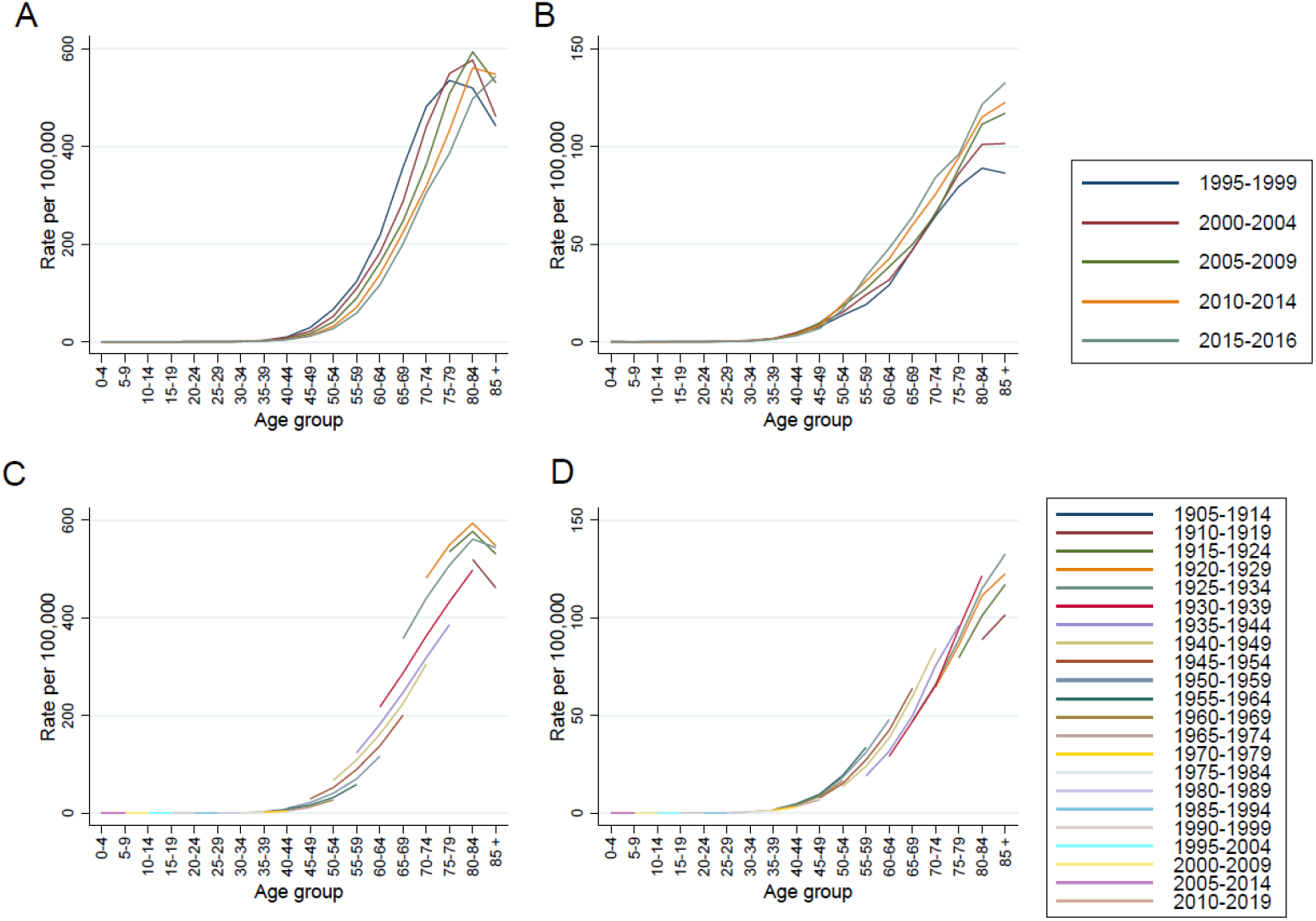
Curves of age-specific lung cancer mortality rates by calendar period (panel A, men; panel B, women) and birth cohort (panel C, men; panel D, women). Italy, 1995-2016

With regard to the SMRs by calendar period, Italy showed a marked decrease among men, while the risk for women was still increasing (Table 2). To better understand these opposite gradients, the age-specific rate curves by calendar period and birth cohort should first be considered. The cross-sectional curves of age-specific mortality rates by calendar period appear to have different shapes (Figure 1). For men, the more recent the calendar period the younger the age group with the highest rate whereas, for women, the more recent the calendar period the older the age group with the highest rate. In other words, the rank of each calendar period depended on which age group we considered, both among men and women. Consequently, the comparison between calendar periods was challenging to summarise and age acted as an effect modifier.

**TABLE 2.** Observed and expected number of lung cancer deaths and age-standardised lung cancer mortality ratios (SMR per 100) and confidence intervals (CI 90%), by sex and calendar period. Italy, 1995-2016

| Calendar period | Men |  |  | Women |  |  |
| --- | --- | --- | --- | --- | --- | --- |
|  | Observed | Expected | SMR (90% CI) | Observed | Expected | SMR (90% CI) |
| 1995-1999 | 128368 | 107149 | 120 (119-120) | 26892 | 31492 | 85 (84-86) |
| 2000-2004 | 127672 | 117046 | 109 (108-110) | 31755 | 34034 | 93 (92-94) |
| 2005-2009 | 126601 | 128896 | 98 (98-99) | 37241 | 36829 | 101 (100-102) |
| 2010-2014 | 123925 | 141288 | 88 (87-88) | 43470 | 39614 | 110 (109-111) |
| 2015-2016 | 48263 | 60450 | 80 (79-81) | 19261 | 16652 | 116 (114-117) |

Abbreviations: CI, confidence interval; SMR, standardised mortality ratio.

On the contrary, the curves of age-specific mortality rates, if viewed longitudinally, i.e. by birth cohort, appeared to be roughly parallel to each other and showed a similar pattern, with higher rates for the same age group in all birth cohorts, i.e. age 80-84 among men and 85+ among women (Figure 1). Furthermore, the rank of each birth cohort was the same in all age groups for men and women. This behaviour was expected based on the natural history of the disease.^30^ The observed data, therefore, supported a modelling approach based on the birth cohort time dimension. To better illustrate the consistency and regularity of these patterns, we plotted in Supplementary Figure 2 the curves of birth cohort-specific mortality rates by age group in both sexes.

### 3.2 Age-period-cohort models of lung cancer mortality rates

Fitting the age-cohort model to the data in Table 1 yielded the birth cohort effects estimates (log rate ratios) shown in Figure 2. We took the 1905-1914 birth cohort as a reference category and, in order to compare the time evolution in men and women, we scaled the women cohort effect to be equal to the men cohort effect for the 1930-1939 birth cohort. In men, the birth cohort at highest risk was the one born between 1920 and 1929, i.e. those people who were aged 15-24 years at the end of World War II. For the cohorts born after 1940, a decrease in mortality was observed. This decline continued until a plateau for the cohorts born after 1970. As far as women are concerned, the risk of dying increased progressively by birth cohort and peaked among those born between 1955 and 1964, i.e. those aged 15-24 years at the end of the seventies. Subsequently, a decrease was observed, which was less rapid than that occurring among men. Lung cancer mortality peaked among women 35 years later than among men.

**FIGURE 2.**
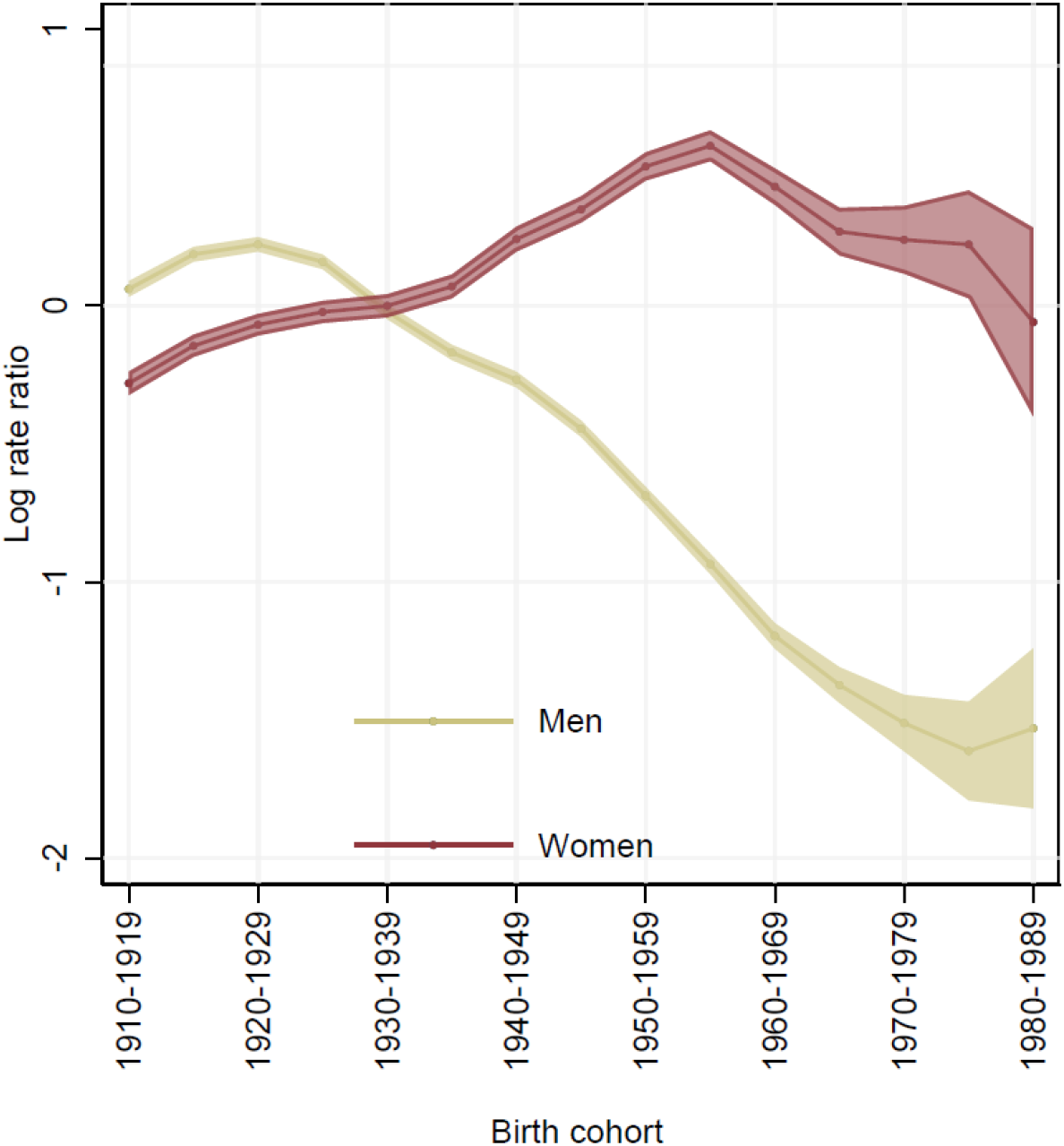
Cohort effects estimates from the age-cohort model fitted on lung cancer mortality rates by sex (yellow lines depict men, red lines depict women) and 90% confidence interval. The cohort effects estimates (log rate ratios) were obtained taking the birth cohort 1905-1914 as a reference and scaling the women cohort effect to be equal to the men cohort effect for the 1930-1939 birth cohort. Italy, 1995-2016

We also fitted a series of age-period-cohort models to the whole national data to check for the presence of a non-linear calendar period effect above the birth cohort effect. We did not find evidence for this (women: Likelihood Ratio chi-square test 4.40, *P*-value = 0.22 comparing the age-period-cohort model versus the age-cohort model; men: Likelihood Ratio chi-square test 0.14, *P*-value = 0.98, comparing the age-period model versus the age-drift model and adjusting for overdispersion) (data not shown).

### 3.3 Bayesian space-time modelling of lung cancer mortality rates by Italian provinces

The time evolution of death rates was not homogeneous. We specified a Bayesian space-time model to study geographic variability of time patterns. The rationale of the Bayesian inference was mainly driven by the sparsity of data when considering the set of Lexis diagrams, one for each of the 107 Italian provinces.

Figure 3 (panel A: men; panel B: women) shows the geographic pattern of the RR in men and women by the nine birth cohorts 1920-1929, …, 1960-1969 predicted by the Bayesian space-time model. Among men, in all Italian provinces, we observed a clear-cut reduction of the risk of dying starting with the earlier birth cohort. The range of the RRs was smaller for the younger cohorts compared with the older ones, which indicates a more homogenous geographic pattern over time. A stronger spatial pattern was evident for the older birth cohorts, with a higher risk in the north and a lower risk in the south. Among women, we observed a rise in RRs peaking for the 1955-1964 birth cohort, but with some variability by province. A north-south gradient and a tendency to decrease differences among provinces were much less evident than in men. In both sexes, the north-south decreasing mortality gradient has been replaced by a west-east decreasing gradient penalising the provinces facing the Tyrrhenian sea.

**FIGURE 3.**
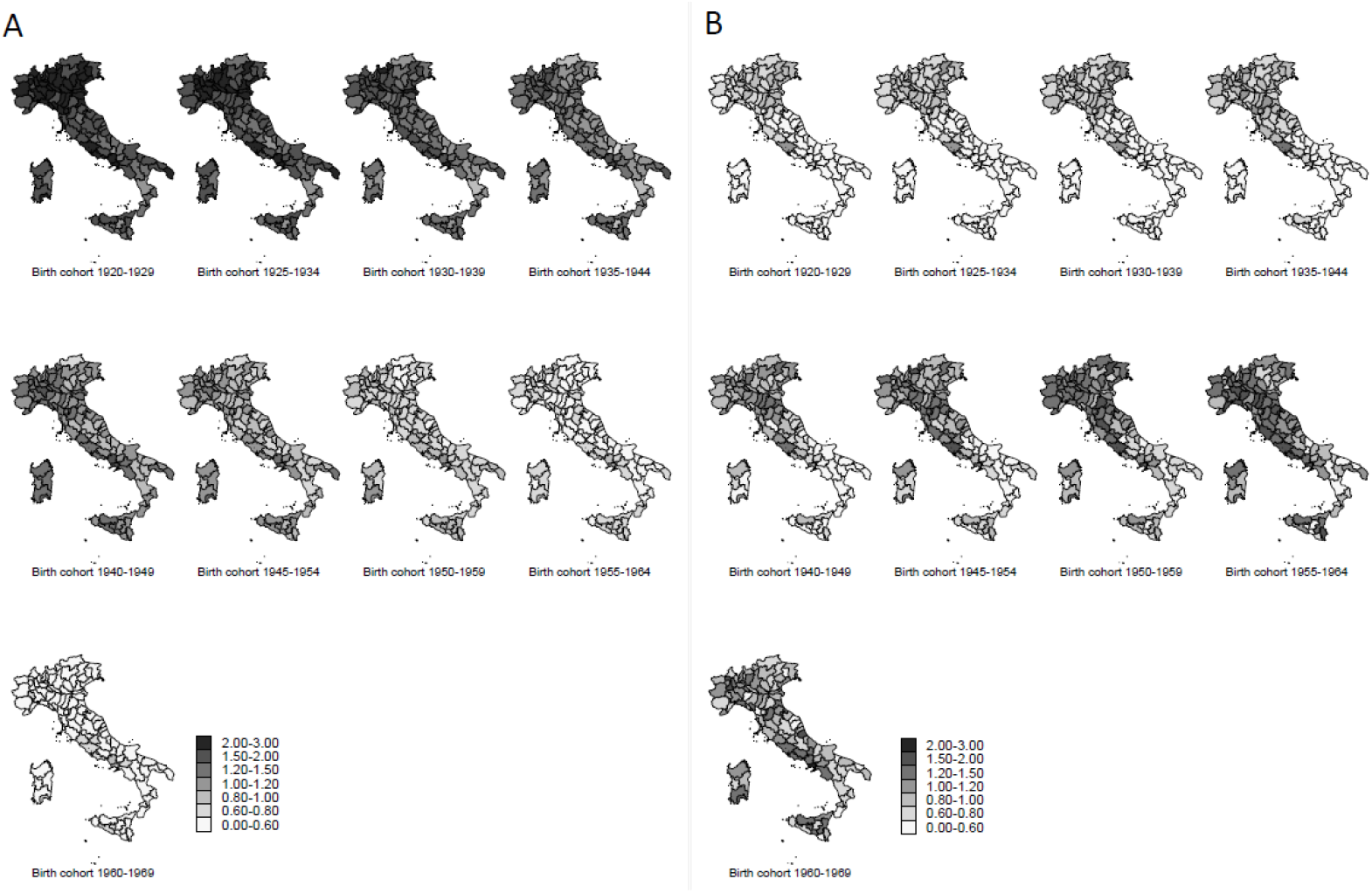
Geographic distribution of lung cancer mortality Bayesian relative risks by province. Absolute scale. Panel A: men; panel B: women. Italy, 1995-2016

Figure 3 (panels A and B) depicts the maps of RRs by sex on an absolute scale. Supplementary Figure 3 and Supplementary Figure 4 depict the maps of RRs by sex on a relative scale. The nine birth cohort-specific Spearman correlation coefficients between the series of provincial RRs observed in men and women ranged between 0.32 and 0.80 (Supplementary Table 1). Supplementary Figure 5 and Supplementary Figure 6 depict the maps of SMRs by sex on an absolute scale. Finally, Supplementary Figure 7 and Supplementary Figure 8 depict the maps of SMRs by sex on a relative scale.

In Figure 4, the birth cohort trends by province are shown and the risk profiles of some selected provinces are highlighted. The reduction and the tendency to geographic homogeneity are clearly depicted in men. However, there was a large difference in the rate of decrease by province. In north-eastern provinces –for example, the province of Venice, in red– the drop was very steep and their ranks changed substantially, as they moved from the higher positions for the 1920-1929 birth cohort to the lower positions for the 1960-1969 birth cohort. In southern provinces (for example, the province of Palermo in Sicily, in blue) the rate of decrease was smoother in the older cohorts. The province of Naples (in purple) was outlying, since it had the highest risk in all birth cohorts, and its gap with the rest of Italy did not show any sign of slowing down. In Supplementary Figures 9 to 19, the birth cohort trends by province in the main Italian administrative regions are shown.

**FIGURE 4.**
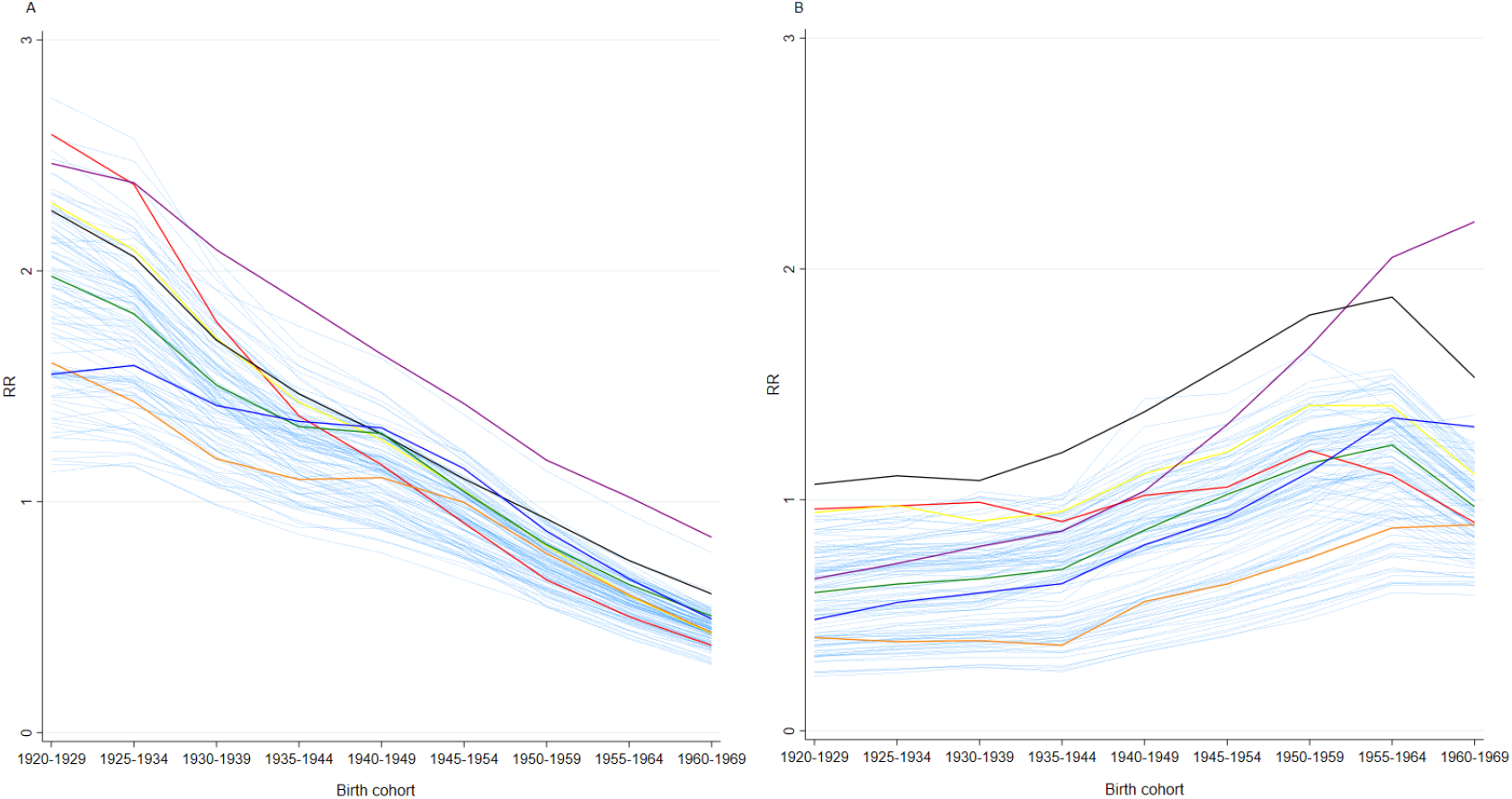
Lung cancer mortality Bayesian relative risks (RR) by birth cohort and province (panel A, men; panel B, women). Highlighted are Venice: red; Milan: yellow; Naples: purple; Rome: black; Prato: green; Oristano: orange; Palermo: blue. Italy, 1995-2016

For women, the trends in Figure 4 differed widely by province. A tendency to geographic homogeneity was evident only for the youngest birth cohort (1960-1969). In the province of Venice (in red), the oldest birth cohort ranked around the maximum level of risk while the youngest one ranked in the lowest quartile. The risk reduction started with the 1950-1959 birth cohort, a pattern consistent with all the north-eastern provinces. Moving from the north-east to the south, the birth cohort with the highest rate changed, being the 1955-1964 cohort for north-central Italy and the 1960-1969 cohort for southern Italy. The rate of decrease was smoother in the southern provinces. Some of them have not reached the peak yet, notably in Campania and Sardinia. The province of Naples was also outlying for women. The risk became the highest at the national level since the 1955-1964 birth cohort, the cohort for which the level of risk exceeded the one seen in women of the province of Rome.

Comparing women with men at the provincial level, the two mortality peaks appeared to be separated by 30 years in Venice, 40 years in Rome, and at least 50 in Naples, where no reversal of the trend is discernible yet.

## 4 Discussion

### 4.1 Temporal trends in the context of European data

The above findings can be better interpreted by contextualising them in lung cancer mortality trends reported elsewhere in Europe. Among men, mortality rates started to decrease –for example– in the late 1970s in Sweden, Finland and the U.K., during the 1980s in the Netherlands, Belgium and Switzerland, and in the late 1990s in Spain and several eastern European countries.^6^ This suggests a trend towards decreasing mortality that has spread from north-western to south-eastern Europe. Since its inception, the decrease appeared to be a birth-cohort-related phenomenon. In Sweden, Finland and the U.K., for example, the downtrend was first observed in those cohorts born in the first decade of the 20th century, whereas in southern Europe the earliest cohorts involved were those born after World War II. In any case, the decrease involved all subsequent cohorts, replacing the previous high-risk ones, which reduced overall mortality rates.

Among women, lung cancer mortality has reached a plateau or has started to decline in some countries of north-western Europe, particularly in the U.K., Ireland and Iceland. This is due to a generational downtrend in the cohorts born after 1950. Conversely, total rates are still growing in Finland, Sweden and Norway, although a trend reversal is expected for the near future. In contrast, but with some exceptions, the rates are increasing in western Europe (especially in France and The Netherlands) and in most southern European countries up to the cohorts born until the 1960s.^4,6^ Noteworthy, the generations of European women at maximum risk have been observed from five (as in Hungary) to 50 years (as in The Netherlands) after the equivalent male cohorts.^4,6^

These differences suggest that, in many countries, men and women are in different phases of the lung cancer epidemic. Our findings show that this is also the case for Italy. For men, we can confirm previous studies of regional-level mortality, which showed a peak in the 1920-1929 cohort, i.e. the generation who were young men after World War II, followed by a generalised decline throughout the country. For women, we can formally confirm the circumstantial findings of previous smaller-scale mortality studies.^9,10^ Mortality has peaked in the 1955-1964 cohort, the generation of women who were students during and after the social movements of the late 1960s. Subsequently, a downward trend has begun in many Italian provinces, with more exceptions in southern Italy (Supplementary Figures 7 and 13-15). This is an original finding, because previous regional-level data had shown a generalised increase even in the most recent birth cohorts studied.^10,31^

### 4.2 Spatial trends

The data reported here also demonstrate that the time trend in lung cancer mortality by sex is interrelated with a geographic transition, i.e. a change in the geographic pattern of rates. The long-standing north-south decreasing mortality gradient has been replaced by a west-east decreasing gradient, with a drop of rates in the regions facing the Adriatic Sea and a different situation in those facing the Tyrrhenian Sea, where mortality RR is stagnating or still worsening. Thus, we can confirm the consistency of a previous observation showing, for the first time, a decline in mortality in the high-risk regions of north-eastern Italy paralleled by an opposite change in the Campania Region.^10^ Among men, the provinces along the Tyrrhenian Sea now rank first in mortality, and, in particular, Naples has taken the place of Venice as the province at highest risk. For women, the change in mortality ranking is consistent with that seen among men, with a decreased risk in northern and north-eastern provinces for the cohorts born in the 1950s and after and in central Italy for the cohorts born since the 1960s. In southern Italy, no cohort-dependent reduction has occurred yet. As for men, the highest risk was in the province of Naples.

The analysis of spatial trends deserves two more comments. First, provincial-level data offer several other pieces of information that are beyond our scope but may be interesting from a local perspective. In general, they result from socio-economic peculiarities. For example, the province of Carbonia-Iglesias showed higher RR among men compared to the other Sardinian provinces, depending on the occupational exposure in the mining industry.^31^ In the Lombardy Region, the mortality drop for women occurred earlier in the province of Como, the one with the highest socio-economic standard. In the Veneto Region, among men, mortality remained notably higher in the province of Rovigo, a socio-economically ‘frail area’.^32^

Second, mortality trends at the provincial level are influenced by the trends occurring in capital cities and major cities. Therefore, the evolution of RR for rural and sparsely inhabited municipalities could not be well described by our data, especially in southern Italy and particularly so among women.

### 4.3 Changing health issues and inequalities

This multifaceted epidemiologic passage, consisting of a temporal and a spatial component, has never been previously reported in Italy. Over the decades, the highest levels of exposure to risk factors for lung cancer have migrated across the country. This changeover poses two problems. The first is that the maximum mortality risk is moving to geographic areas where 5-year age-standardised net survival (men, 13%; women, 18%) is lower than in the rest of the country (north-eastern Italy: men, 16%; women, 20%. North-western and central Italy: men, 15%; women, 19%).^33^ The second problem is that the design of smoking prevention and cessation campaigns needs to be reconsidered, in order to determine whether they should be tailored to a changing target population.^4^

A third correlate of the transition is that the variability of provincial mortality rates has changed in opposite directions for men and women. In the earliest cohort, the heterogeneity of rates was greater for men. In the most recent one, the opposite is seen. In fact, this is mostly accounted for by the huge mortality increase observed among women living in Naples, specifically among those born after World War II. This subset of the Italian female population warrants an in-depth evaluation.

### 4.4 Factors involved in temporal and spatial trends

The temporal and spatial patterns by which the upward trend in lung cancer mortality ceased in Europe, including Italy, among men and the different patterns observed among women suggest that different factors were involved. For men, the reversal of the trend has spread from north-western Europe to south-eastern Europe, which suggests that an improving socio-economic status has been the major driver of the change in smoking habits. This would be consistent with the known inverse association between multiple socio-economic status indicators and disease risk.^34^ In Italy, the changing direction of the geographic mortality gradient mirrors the fact that the regions facing the Adriatic Sea –more specifically, Marche, Abruzzo, and Puglia– have attained a high level of economic development several decades after north-western Italy.^35-37^

As regards women, the socio-economic gradients have certainly had a role in determining the previous north-south and the current west-east mortality gradients. Conversely, the socio-economic factors cannot explain the substantial delay with which the reversal of the mortality trend occurred. We raise the hypothesis that Italian women have modified their smoking habits about 35 years after men because they had a much lower baseline risk. Consequently, the upward mortality trend has caused a sufficient level of social alarm much later among women. This view is supported by the consideration that the attitudes of Italian women toward another cancer epidemic, that of melanoma, have been definitely different. With a roughly comparable incidence of disease,^38^ women have preceded men in adopting sun avoidance and skin self-surveillance practices.^38,39^

It is worth considering that the socio-economic gradients and the lower baseline risk of lung cancer have probably cooperated in influencing women’s attitudes towards smoking. This would explain why the delay of the mortality peak for women versus men was 30 years in Venice, 40 years in Rome, and at least 50 in Naples, where the trend for women is still increasing.

### 4.5 Strengths and weaknesses

Investigating the birth cohort effects allowed us to anticipate future mortality rates, because these will be related to current mortality trends in the most recent generations. We emphasise that the analysis of trends by birth cohort encompasses a wide time span and enables describing the evolution of mortality in a longer time perspective than the one allowed by cross-sectional analyses.

This study has a high degree of novelty for three reasons. First, and most important, Italian mortality data for the years 1995-2016 have never been previously reported.

Second, we broke down the analysis into as many as 107 provinces, which is an opportunity so far seldom available to studies of trends by birth cohort in Europe and beyond.^12,13^ A province in Italy is a small geographic unit. The small geographic basis of the data, coupled with great statistical power, made it possible to obtain a high-resolution overview of the multifaceted changes that are ongoing.

And third, data were derived from the national mortality registry. As a result, this study was free of the selection bias potentially affecting incidence studies from those countries where cancer registration covers only part –not always constant over time– of the national population. This is the case, among others, for Italy, where most of the currently available incidence data are from the northern regions.^38,40^

There are also methodological issues in this study that need to be pointed out. First, the age group and the calendar period did not always have the same dimension due to limitations in data availability. However, this problem was restricted to the last time period and only impacted the last birth cohort studied.

Second, different specifications are possible for the space-time interaction term. In our model, the interaction terms were structured both in space and time.

Third, we used data cross-classified in 5×5-years cells in the Lexis diagram with age groups spanning from 0-4 to 85+ years. Because of the Italian population’s ageing, it is common to find in current literature that the last age group is splitted into three further age groups (85-89, 90-94, 95+). We did not do it in order to be consistent with the population structure in the nineties (the percentage of people of 85 years was among men and women respectively 0.5% and 1.2% in 1995, 0.6% and 1.5% in 2000, 0.6% and 1.4% in 2005, 0.8% and 1.9% in 2010, 1.0% and 2.2% in 2015, 1.2% and 2.4% in 2020). The last calendar period studied was 2015-2016, a two-year period, but we assumed the mortality estimates for this period to be valid for the whole period 2015-2019. In the birth-cohort dimension, our assumption was similar. For example, the 1960-1969 cohort was observed in the age group 50-52, but we assumed the estimates to be valid for the age group 50-54. However, major biases are unlikely, because the number of cases in these cells was low.

Furthermore, this study might be affected by differences or temporal changes in access to effective diagnosis and treatment techniques. However, these cannot necessarily be considered to introduce a bias into results. An increased healthcare expenditure, with improved quality and accessibility of services, is a correlate of economic growth, thus reinforcing the inverse association of socio-economic indicators with lung cancer mortality.

### 4.6 Conclusions

This study is a substantial update to previous knowledge on lung cancer mortality in Italy. We conclude the following: (1) among men, mortality was confirmed to have reached a peak in the 1920-1929 cohort, followed by a generalised decline; among women, there was novel evidence for a peak in the 1955-1964 cohort, followed by an unevenly distributed downward trend; (2) the north-south decreasing gradient has been replaced by a west-east decreasing gradient, with Naples currently being the province at highest risk for both sexes; (3) the diversity of mortality rates was greater for men in the earliest cohort and for women in the most recent one; (4) for both sexes, an improving socio-economic status has been the major driver of the change in smoking habits, with an average 35-year delay being observed for women; and (5) the delay, in fact, was 30 years in Venice, 40 years in Rome, and at least 50 in Naples, where the trend for women is still on the rise.

## Data Availability

Research data are available from Giada Minelli upon reasonable request.

## ABBREVIATIONS

CI: confidence interval
ICD: International Classification of Diseases
ISTAT: Istituto Nazionale di Statistica
RR: relative risk
SMR: standardised mortality ratio
U.K.: United Kingdom

## AUTHOR CONTRIBUTIONS

**Dolores Catelan:** Conceptualization, Methodology, Validation, Formal Analysis, Writing—Original Draft, Writing—Review & Editing, Supervision. **Annibale Biggeri:** Conceptualization, Methodology, Validation, Formal Analysis, Investigation, Writing—Original Draft, Writing—Review & Editing, Supervision. **Lauro Bucchi:** Investigation, Writing—Original Draft, Writing—Review & Editing, Supervision. **Valerio Manno:** Resources, Data Curation, Writing—Review & Editing. **Marilena Pappagallo:** Resources, Data Curation, Writing—Review & Editing. **Giorgia Stoppa:** Methodology, Software, Validation, Formal Analysis, Data Curation, Writing—Review & Editing, Visualization. **Francesco Grippo:** Resources, Data Curation, Writing—Review & Editing. **Luisa Frova:** Resources, Data Curation, Writing—Review & Editing, Supervision. **Federica Zamagni:** Software, Validation, Formal Analysis, Writing—Original Draft, Writing—Review & Editing, Visualization. **Roberta Crialesi:** Resources, Writing—Review & Editing, Supervision. **Giada Minelli:** Resources, Data Curation, Writing—Review & Editing, Supervision. The work reported in the paper has been performed by the authors, unless clearly specified in the text.

## ETHICS STATEMENT

The study was approved by the Ethics Committee at the Romagna Cancer Institute (ID: IRST100.37).

## DATA AVAILABILITY STATEMENT

Research data are available from Giada Minelli upon reasonable request.

## SUPPLEMENTARY MATERIALS

**SUPPLEMENTARY TABLE 1.** Spearman correlation coefficients and 95% confidence intervals between the series of provincial Bayesian Relative Risks in men and women. Lung cancer mortality. Italy, 1995-2016

| Birth cohort | Spearman rho (95% CI)* |
| --- | --- |
| 1920-1929 | 0.801 (0.709-0.869) |
| 1925-1934 | 0.779 (0.677-0.857) |
| 1930-1939 | 0.690 (0.566-0.788) |
| 1935-1944 | 0.541 (0.385-0.677) |
| 1940-1949 | 0.457 (0.289-0.590) |
| 1945-1954 | 0.376 (0.199-0.536) |
| 1950-1959 | 0.315 (0.131-0.479) |
| 1955-1964 | 0.428 (0.264-0.577) |
| 1960-1969 | 0.571 (0.436-0.688) |

Abbreviations: CI, confidence intervals.
\*Bias corrected bootstrapped 95% CIs (5000 iterations)

**SUPPLEMENTARY FIGURE 1.**
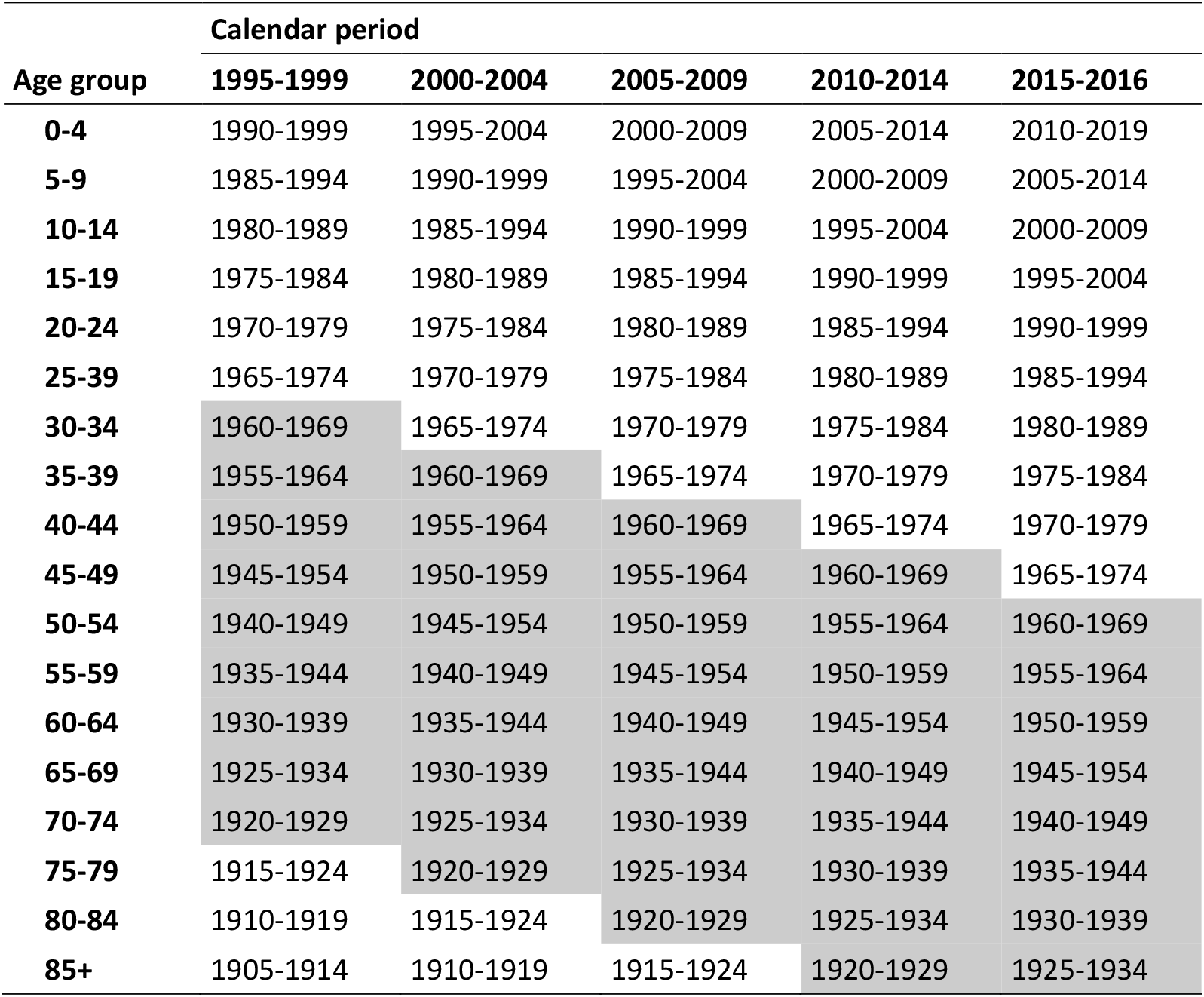
Lexis diagram showing calendar periods, age groups, and birth cohorts. Highlighted in grey are the birth cohorts used in the space-time hierarchical Bayesian model. Italy, 1995-2016

**SUPPLEMENTARY FIGURE 2.**
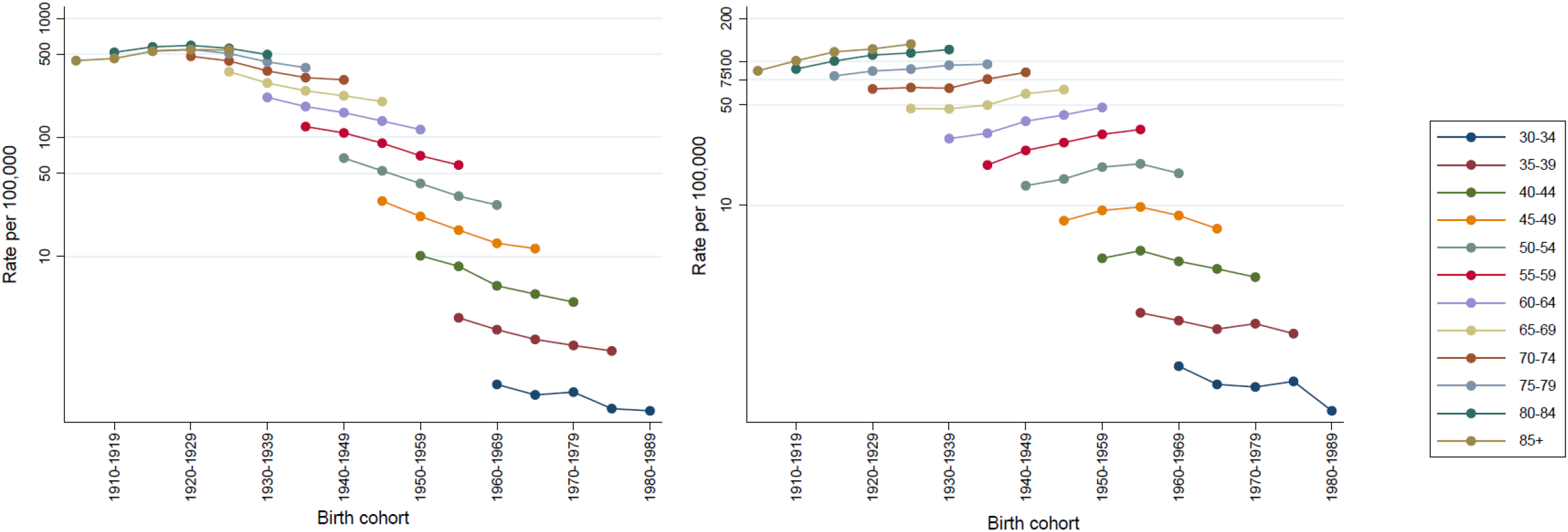
Curves of 5-year age-specific lung cancer mortality rates by birth cohort (panel A, men; panel B, women) displayed on a semi-log scale. From the bottom to the top, the 5-year age groups are 30-34 years, …, 85+ years. Italy, 1995-2016

**SUPPLEMENTARY FIGURE 3.**
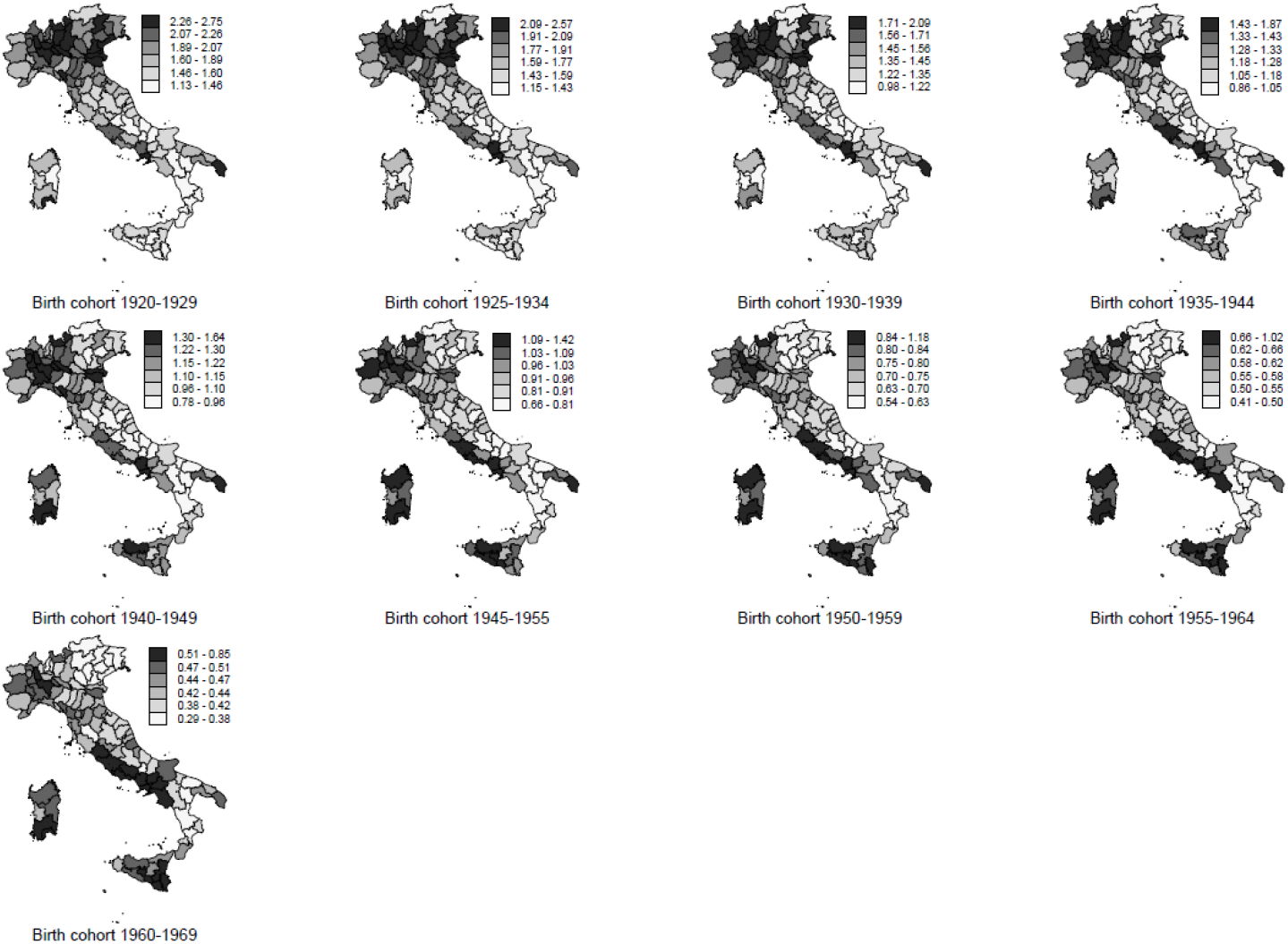
Geographic distribution of lung cancer mortality Bayesian relative risks by province. Relative scale. Men. Italy, 1995-2016

**SUPPLEMENTARY FIGURE 4.**
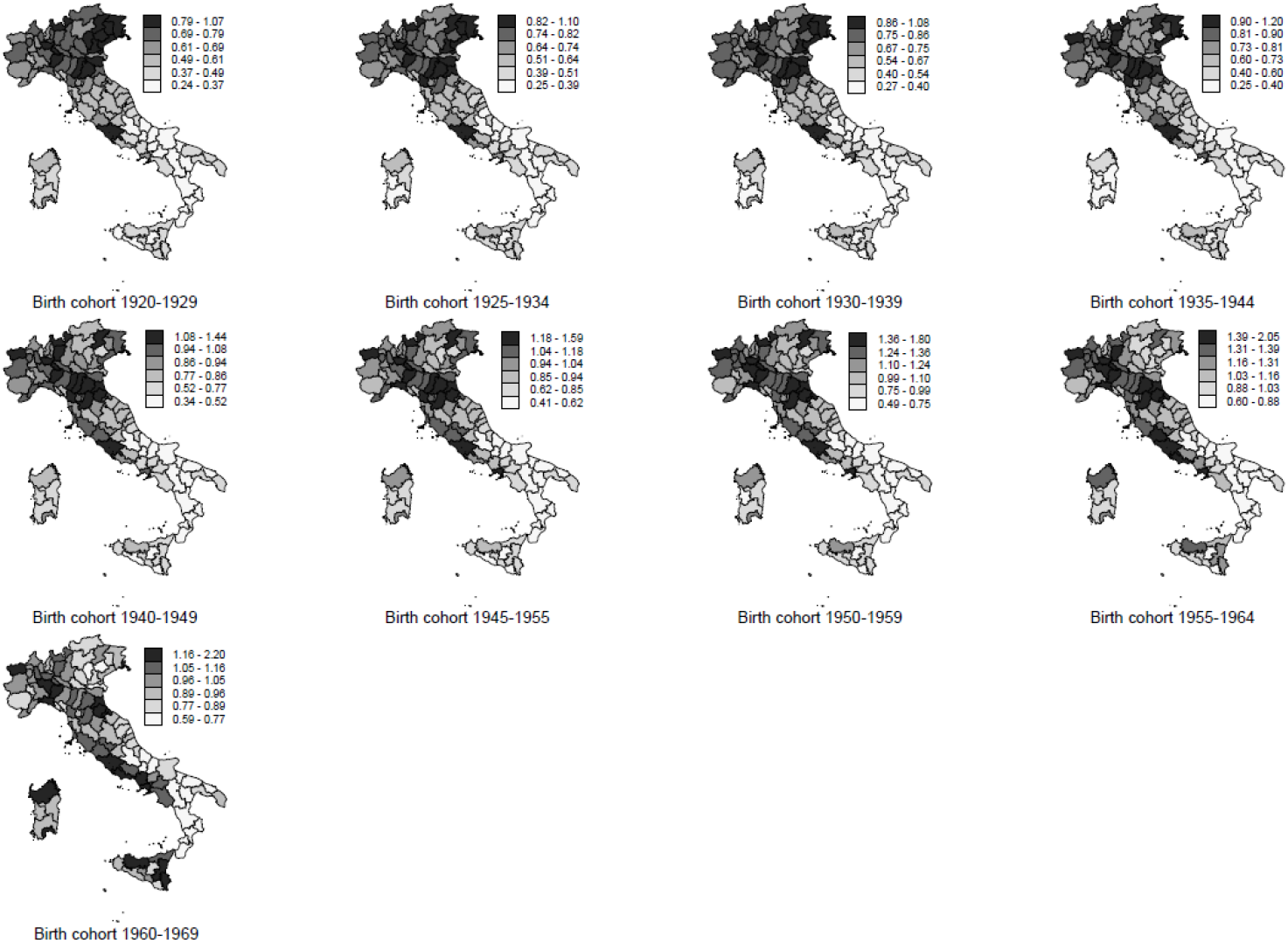
Geographic distribution of lung cancer mortality Bayesian relative risks by province. Relative scale. Women. Italy, 1995-2016

**SUPPLEMENTARY FIGURE 5.**
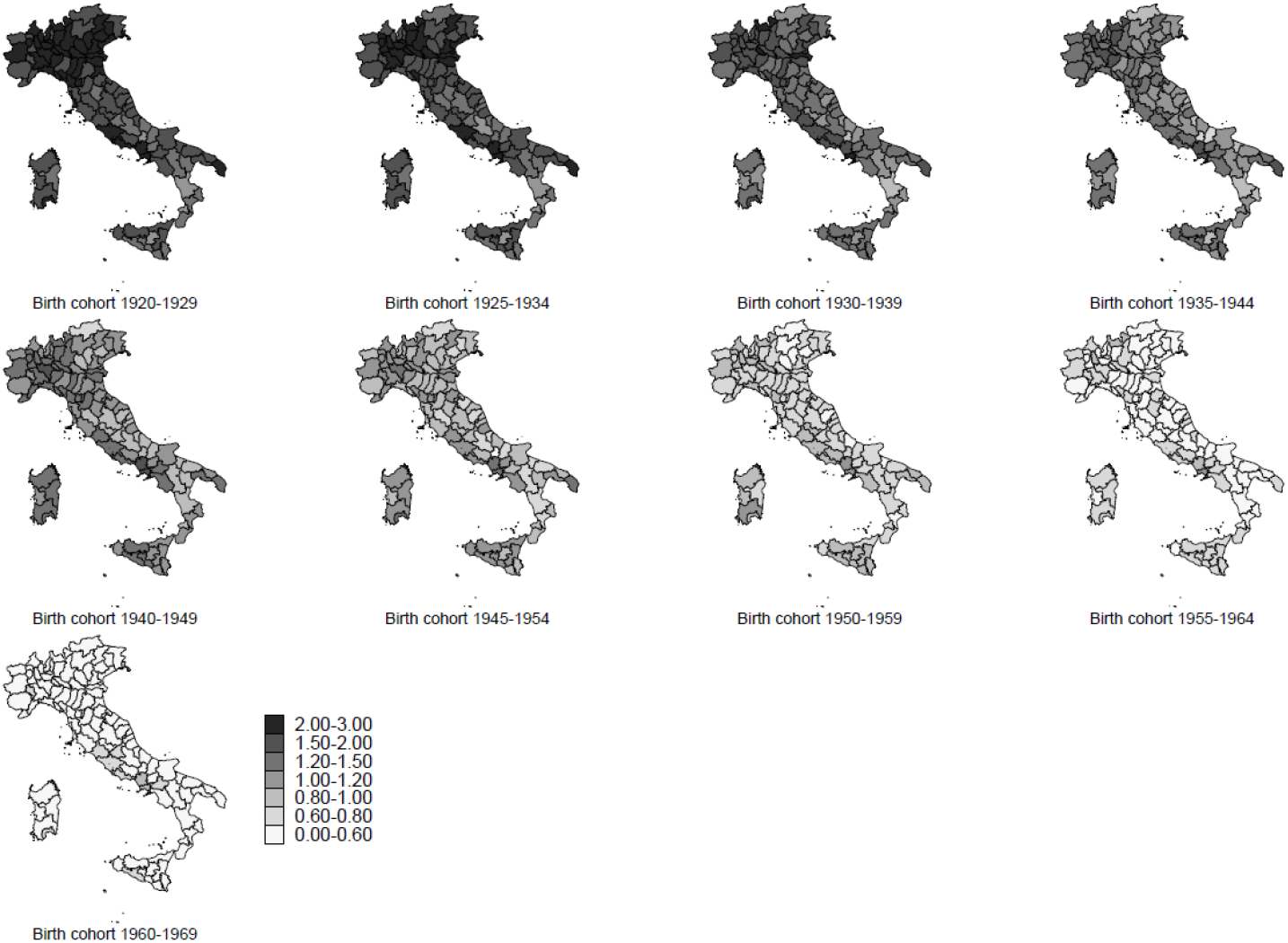
Geographic distribution of lung cancer standardised mortality ratios by province. Absolute scale. Men. Italy, 1995-2016

**SUPPLEMENTARY FIGURE 6.**
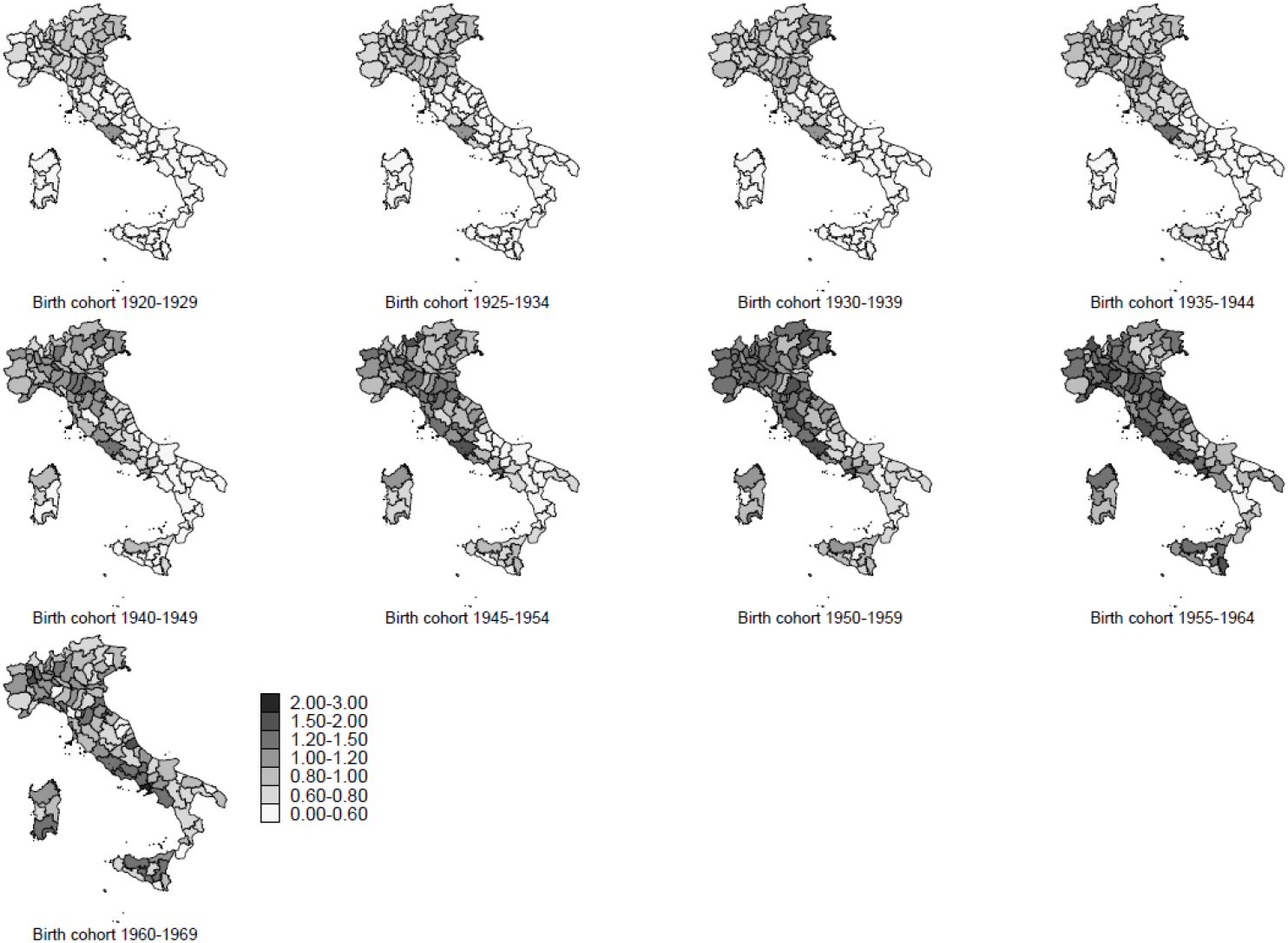
Geographic distribution of lung cancer standardised mortality ratios by province. Absolute scale. Women. Italy, 1995-2016

**SUPPLEMENTARY FIGURE 7.**
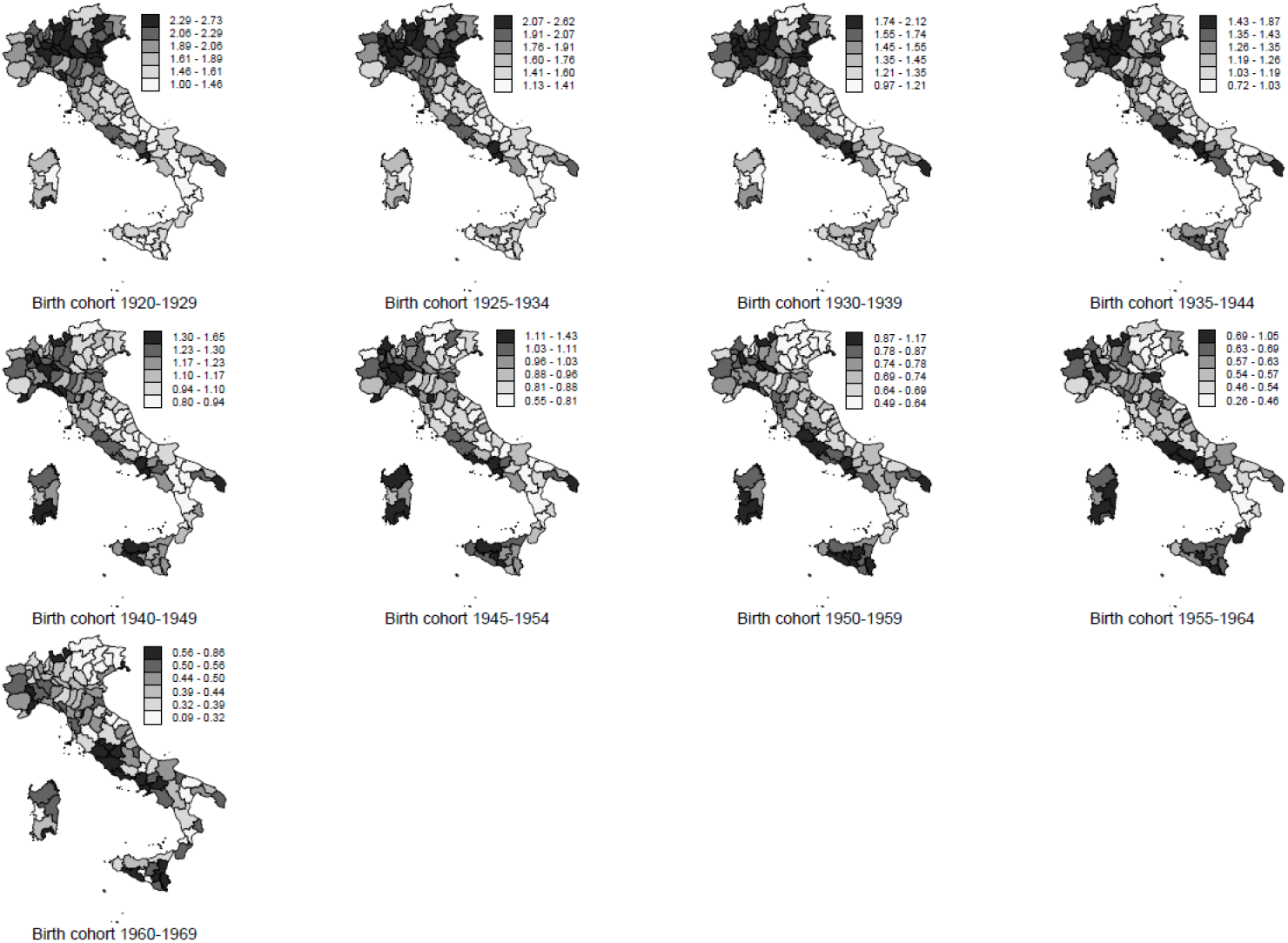
Geographic distribution of lung cancer standardised mortality ratios by province. Relative scale. Men. Italy, 1995-2016

**SUPPLEMENTARY FIGURE 8.**
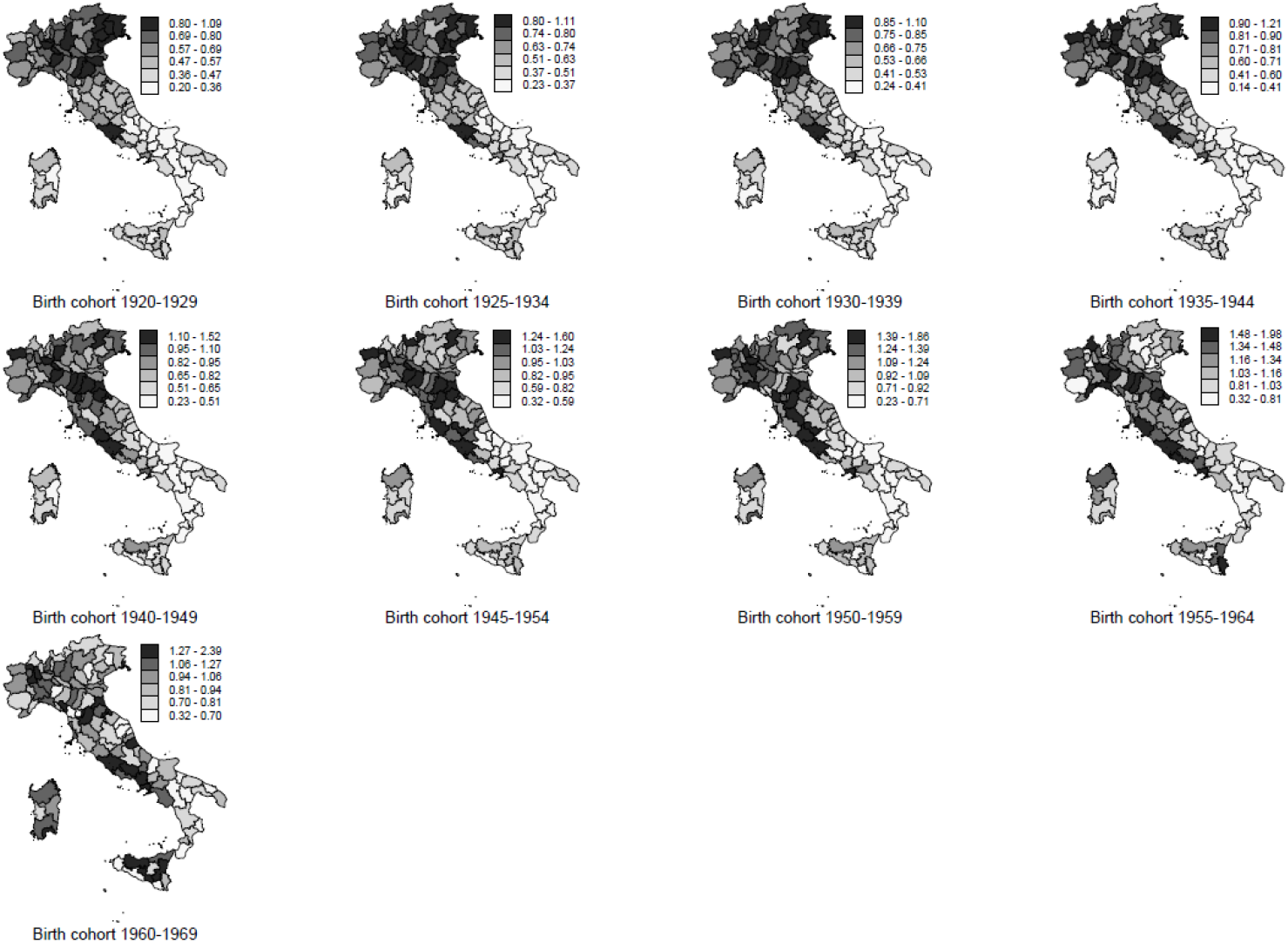
Geographic distribution of lung cancer standardised mortality ratio by province. Relative scale. Women. Italy, 1995-2016

**SUPPLEMENTARY FIGURE 9.**
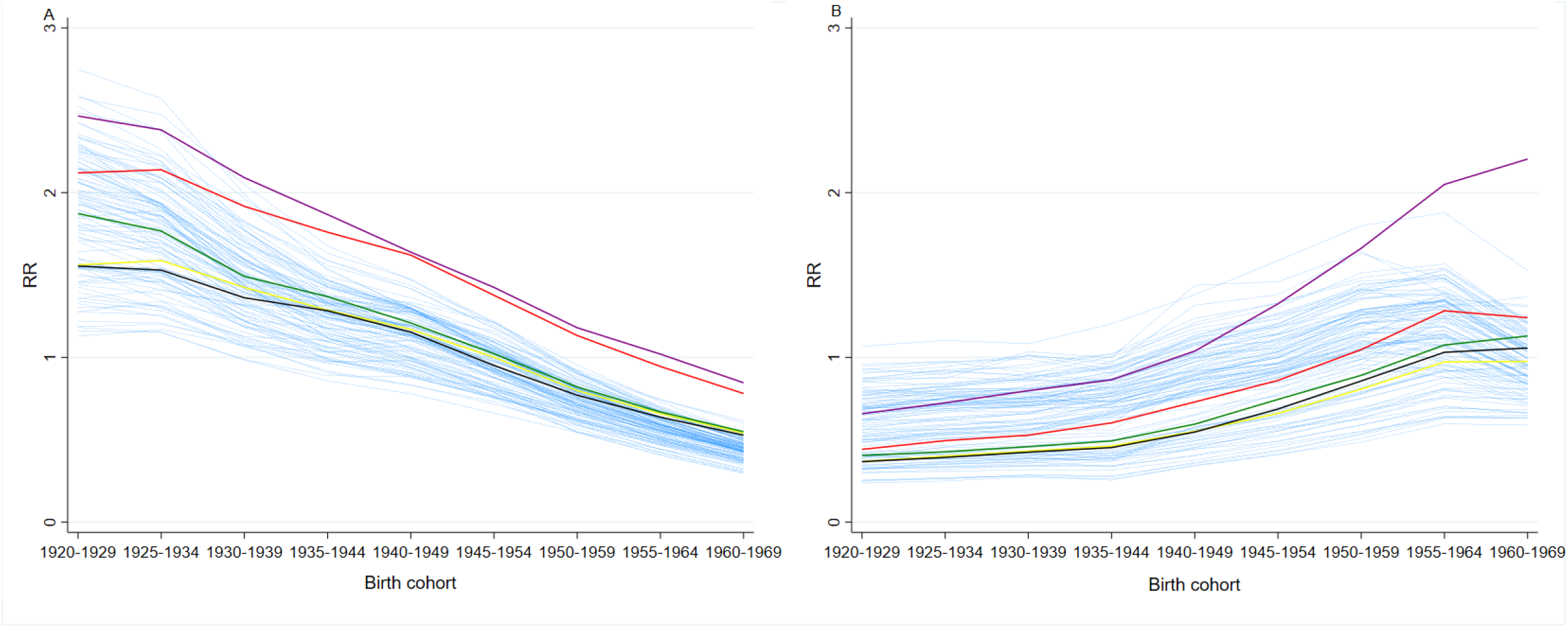
Lung cancer mortality Bayesian relative risks (RR) by birth cohort and province (panel A, men; panel B, women). Campania Region. Highlighted are Salerno: green; Avellino: black; Napoli: purple; Benevento: yellow; Caserta: red; other Italian provinces: light blue. Italy, 1995-2016

**SUPPLEMENTARY FIGURE 10.**
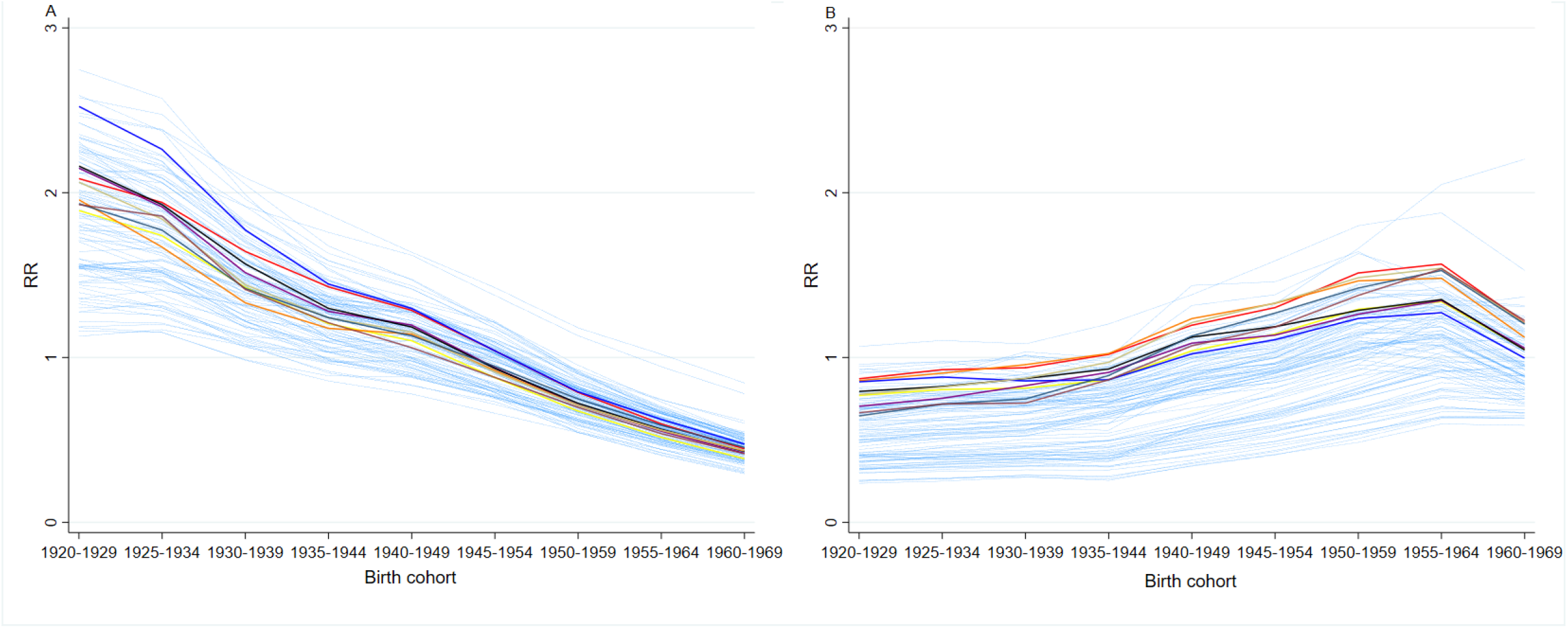
Lung cancer mortality Bayesian relative risks (RR) by birth cohort and province (panel A, men; panel B, women). Emilia-Romagna Region. Highlighted are Rimini: brown; Forlì-Cesena: grey; Ravenna: light green; Ferrara: blue; Bologna: orange; Modena: black; Reggio Emilia: purple; Parma: yellow; Piacenza: red; other Italian provinces: light blue. Italy, 1995-2016

**SUPPLEMENTARY FIGURE 11.**
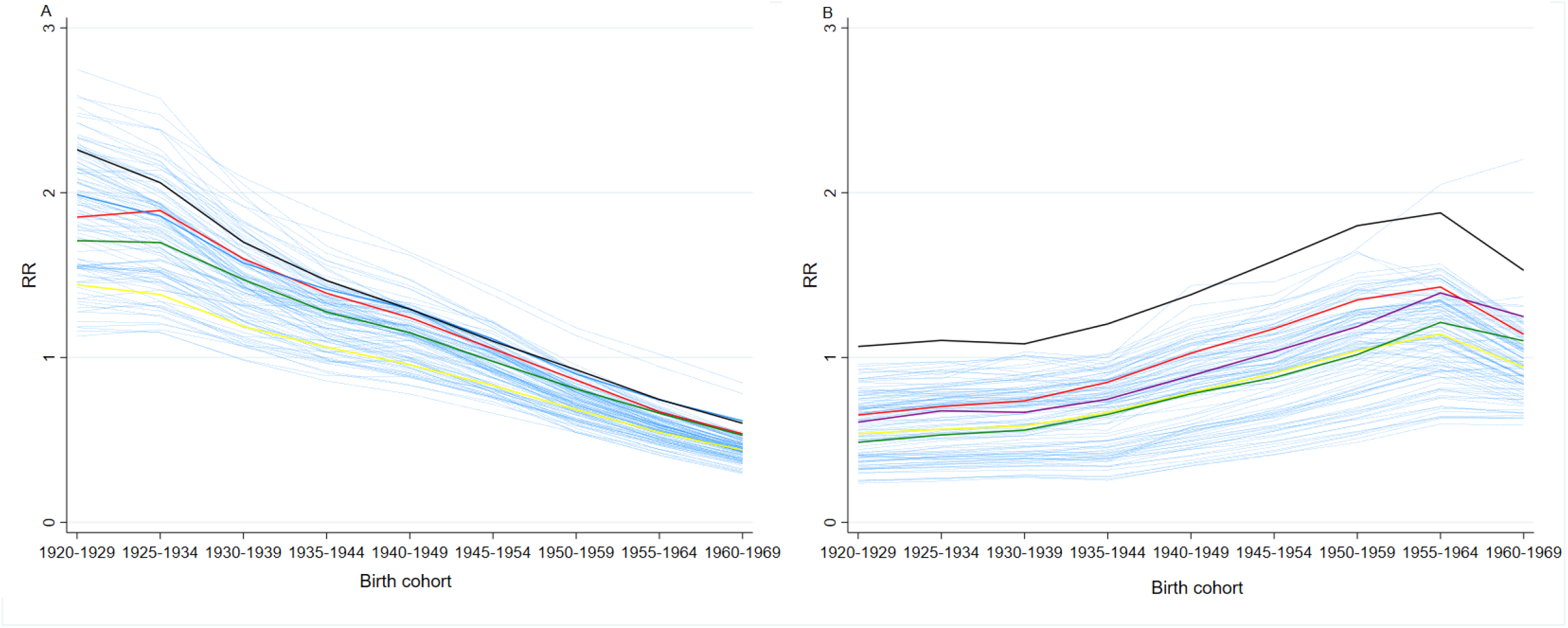
Lung cancer mortality Bayesian relative risks (RR) by birth cohort and province (panel A, men; panel B, women). Lazio Region. Highlighted are Frosinone: green; Roma: black; Latina: purple; Rieti: yellow; Viterbo: red; other Italian provinces: light blue. Italy, 1995-2016

**SUPPLEMENTARY FIGURE 12.**
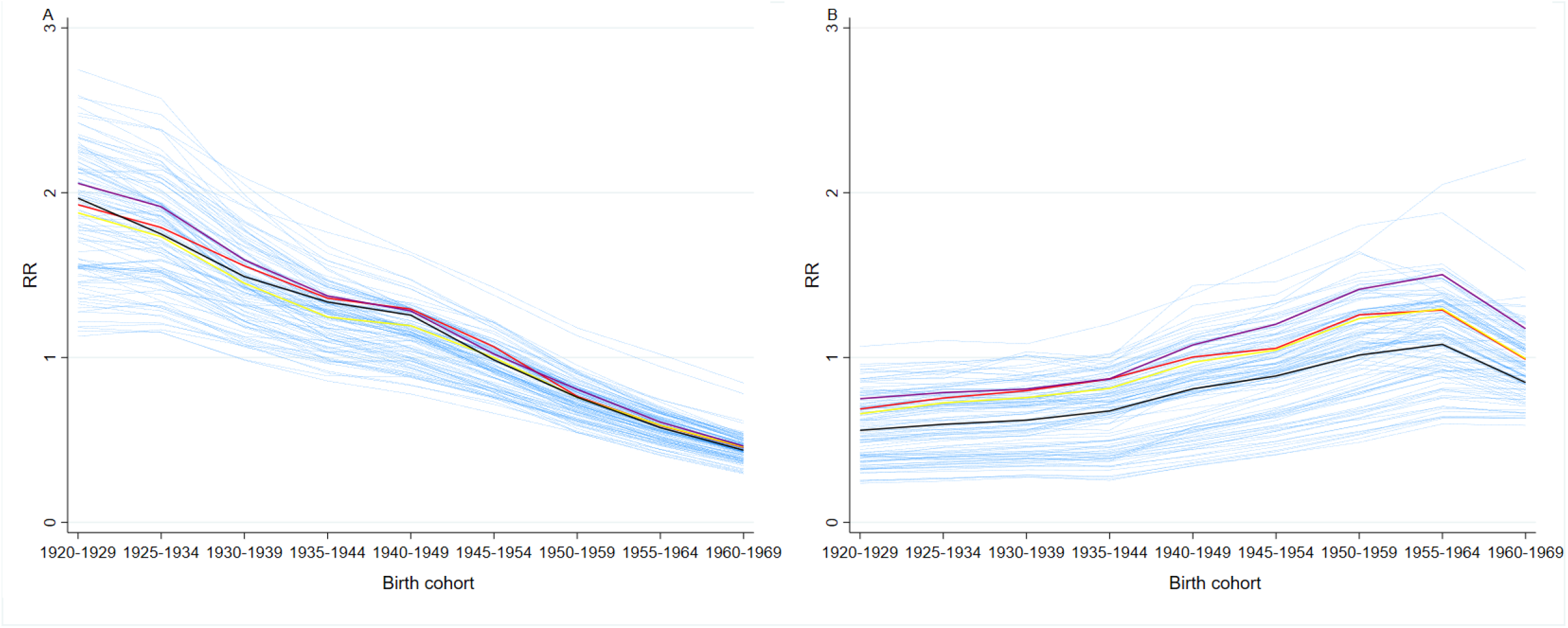
Lung cancer mortality Bayesian relative risks (RR) by birth cohort and province (panel A, men; panel B, women). Liguria Region. Highlighted are La Spezia; black; Genova: purple; Savona: yellow; Imperia: red; other Italian provinces: light blue. Italy, 1995-2016

**SUPPLEMENTARY FIGURE 13.**
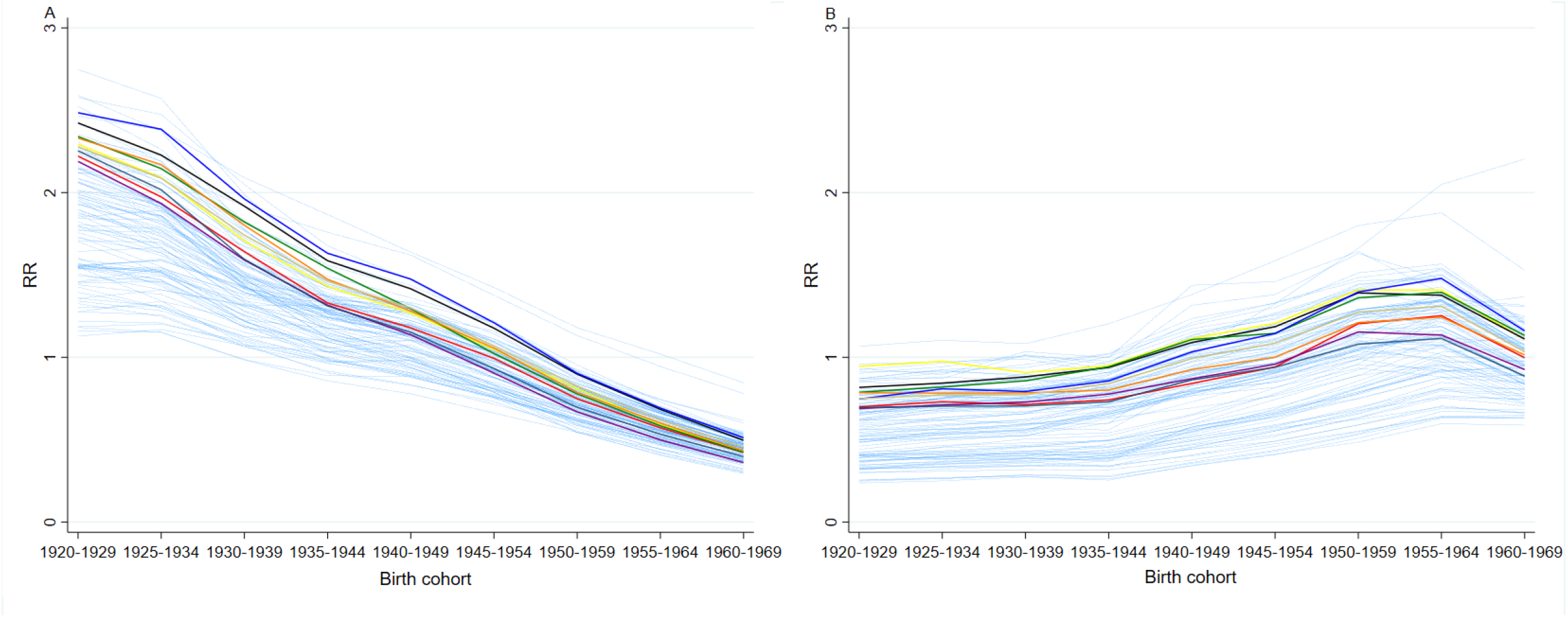
Lung cancer mortality Bayesian relative risks (RR) by birth cohort and province (panel A, men; panel B, women). Lombardia Region. Highlighted are Mantova: grey; Cremona: light green; Pavia: blue; Brescia: orange; Bergamo: green; Sondrio: black; Como: purple; Milano: yellow; Varese: red; other Italian provinces: light blue. Italy, 1995-2016

**SUPPLEMENTARY FIGURE 14.**
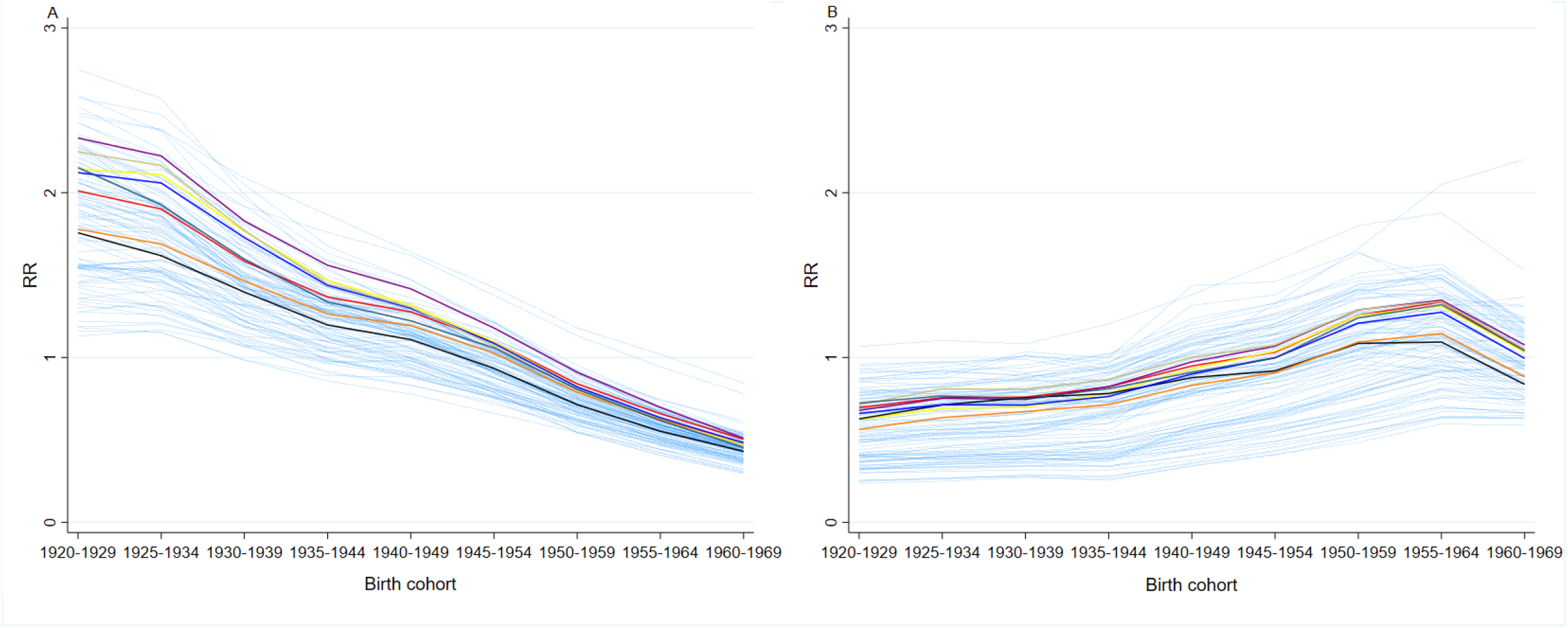
Lung cancer mortality Bayesian relative risks (RR) by birth cohort and province (panel A, men; panel B, women). Piemonte Region. Highlighted are Verbano-Cusio-Ossiola: grey; Biella: light green; Alessandria: blue; Asti: orange; Cuneo: black; Novara: purple; Vercelli: yellow; Torino: red; other Italian provinces: light blue. Italy, 1995-2016

**SUPPLEMENTARY FIGURE 15.**
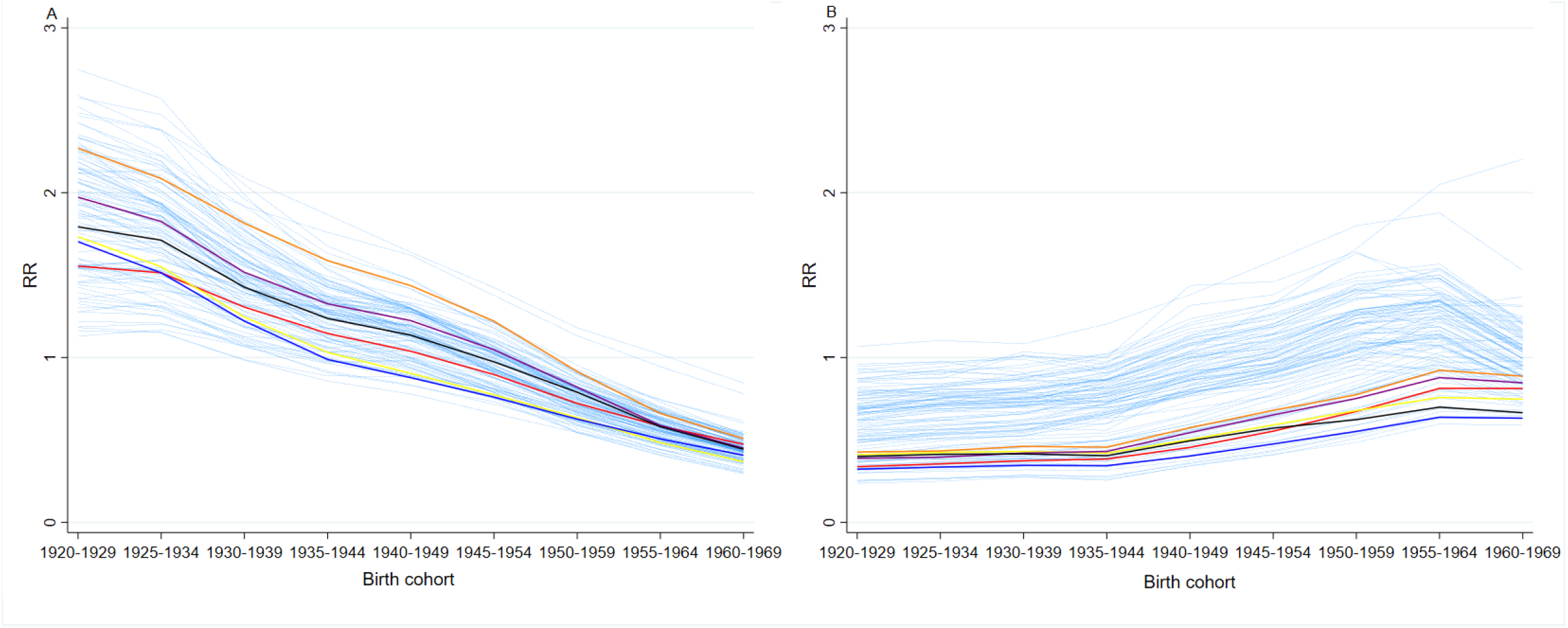
Lung cancer mortality Bayesian relative risks (RR) by birth cohort and province (panel A, men; panel B, women). Puglia Region. Highlighted are Barletta-Andria-Trani: blue; Lecce: orange; Brindisi: black; Taranto: purple; Bari: yellow; Foggia: red; other Italian provinces: light blue. Italy, 1995-2016

**SUPPLEMENTARY FIGURE 16.**
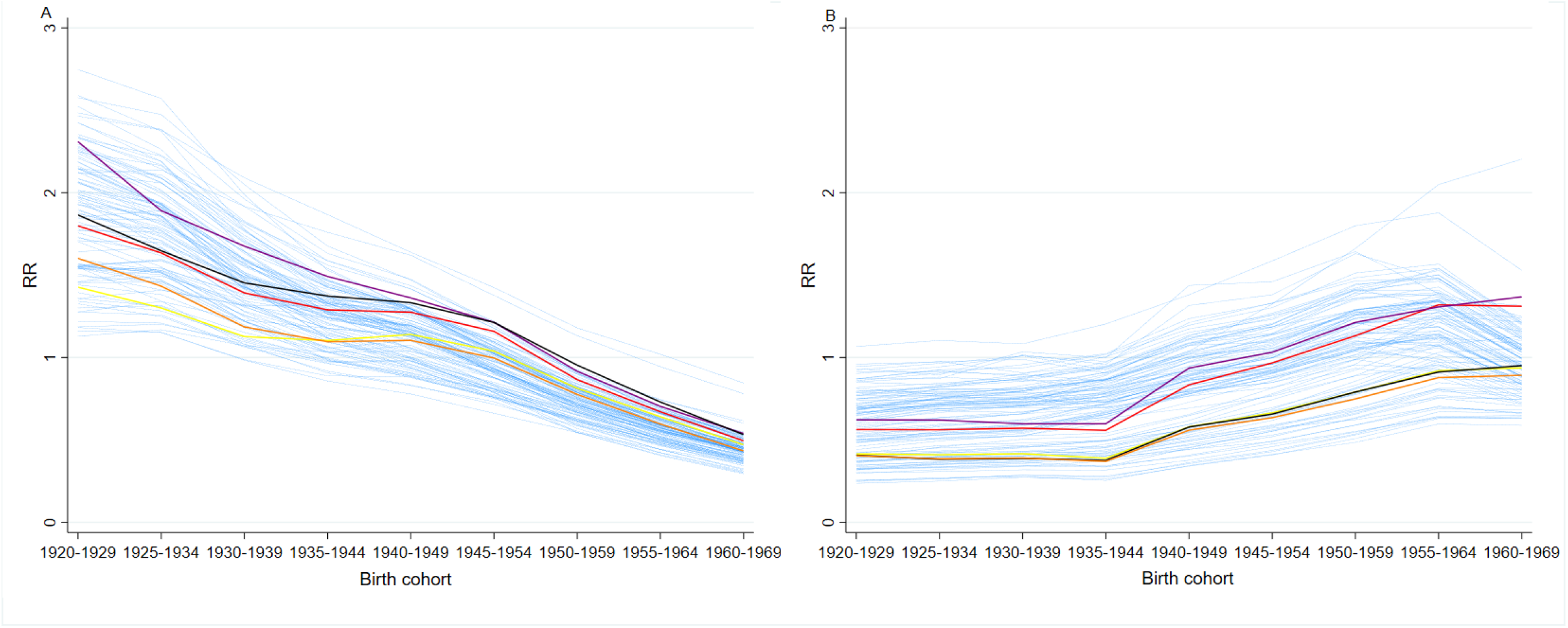
Lung cancer mortality Bayesian relative risks (RR) by birth cohort and province (panel A, men; panel B, women). Sardegna Region. Highlighted are Oristano: orange; Carbonia-Iglesias: black; Cagliari: purple; Nuoro: yellow; Sassari: red; other Italian provinces: light blue. Italy, 1995-2016

**SUPPLEMENTARY FIGURE 17.**
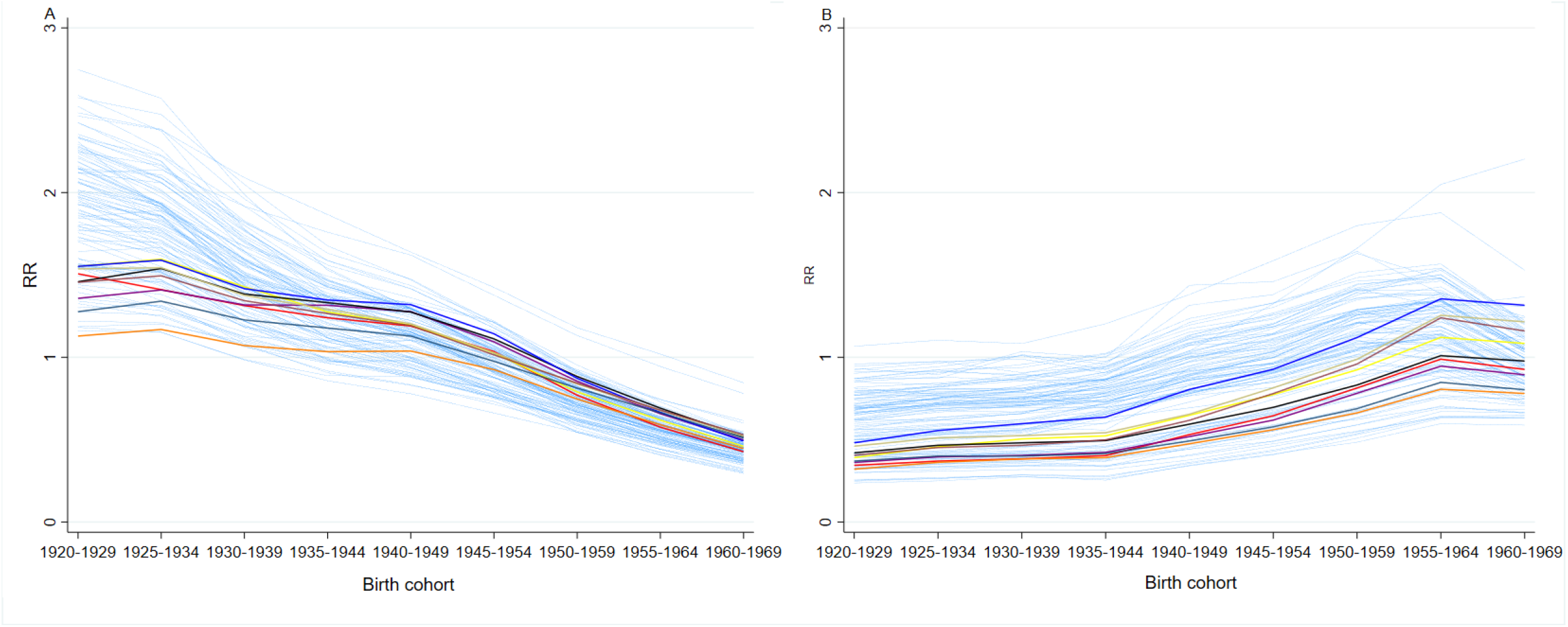
Lung cancer mortality Bayesian relative risks (RR) by birth cohort and province (panel A, men; panel B, women). Sicilia Region. Highlighted are Siracusa: brown; Ragusa: grey; Catania: light green; Palermo: blue; Enna: orange; Caltanissetta: black; Agrigento: purple; Messina: yellow; Trapani: red; other Italian provinces: light blue. Italy, 1995-2016

**SUPPLEMENTARY FIGURE 18.**
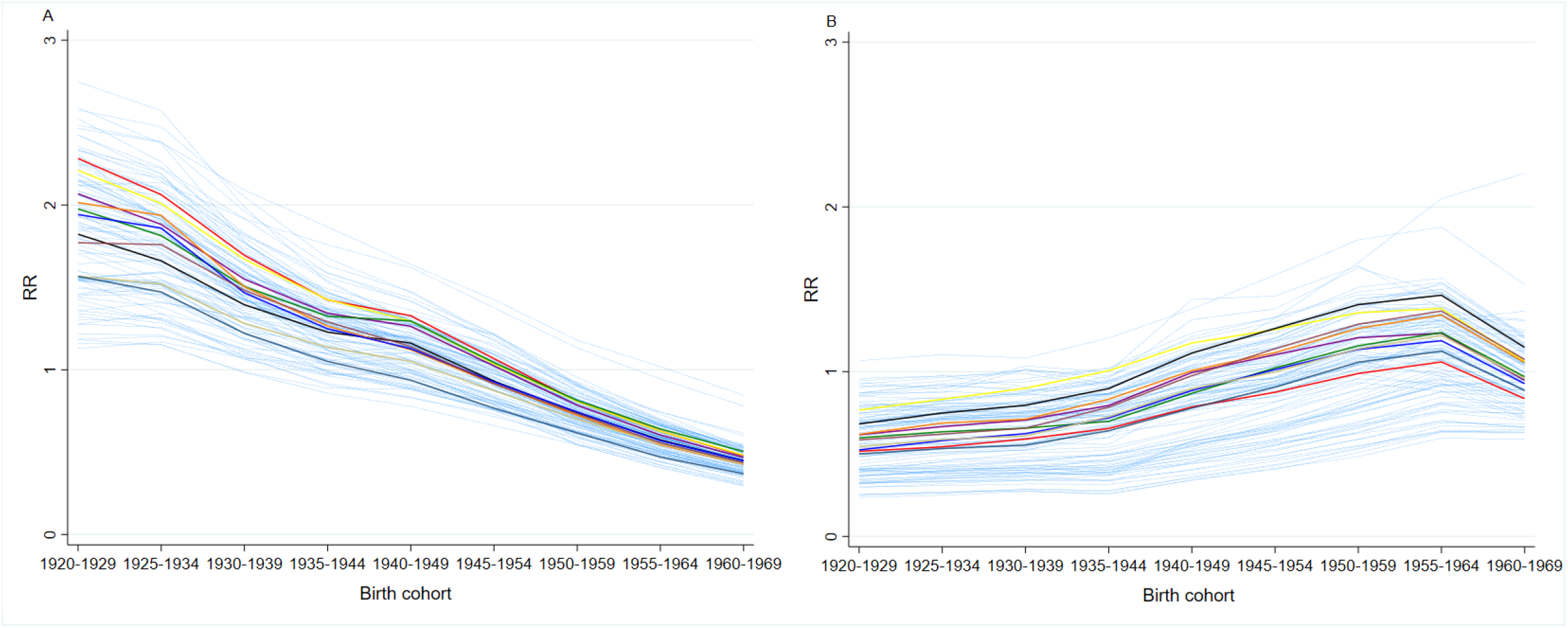
Lung cancer mortality Bayesian relative risks (RR) by birth cohort and province (panel A, men; panel B, women). Toscana Region. Highlighted are Grosseto: brown, Siena: grey; Arezzo: light green; Pisa: blue; Livorno: orange; Prato: green; Firenze: black; Pistoia: purple; Lucca: yellow; Massa Carrara: red; other Italian provinces: light blue. Italy, 1995-2016

**SUPPLEMENTARY FIGURE 19.**
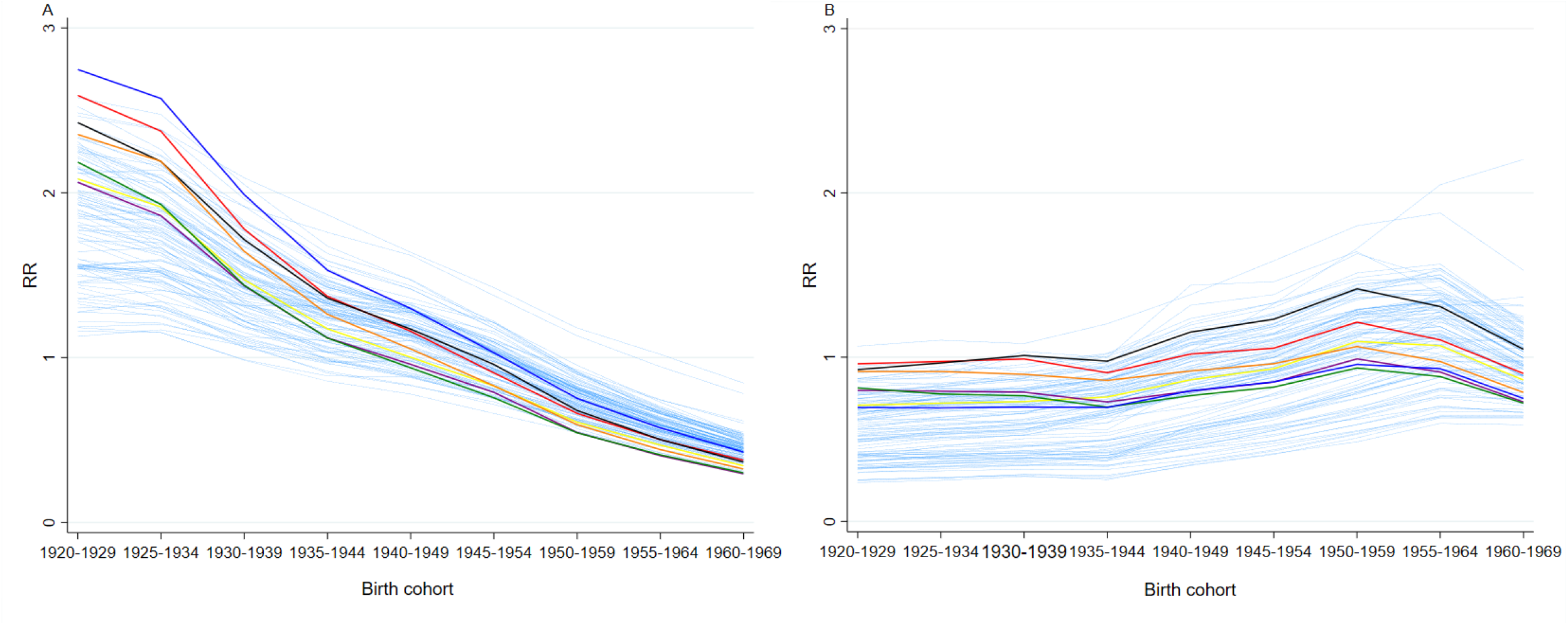
Lung cancer mortality Bayesian relative risks (RR) by birth cohort and province (panel A, men; panel B, women). Veneto Region. Highlighted are Rovigo: blue; Padova: orange; Treviso: green; Belluno: black; Vicenza: purple; Verona: yellow; Venezia: red; other Italian provinces: light blue. Italy, 1995-2016

